# A phase 1/2 randomized, double-blinded, placebo controlled ascending dose trial to assess the safety, tolerability and immunogenicity of ARCT-021 in healthy adults

**DOI:** 10.1101/2021.07.01.21259831

**Authors:** Jenny G. Low, Ruklanthi de Alwis, Shiwei Chen, Shirin Kalimuddin, Yan Shan Leong, Tania Ken Lin Mah, Natalene Yuen, Hwee Cheng Tan, Summer L Zhang, Jean X.Y. Sim, Yvonne F.Z. Chan, Ayesa Syenina, Jia Xin Yee, Eugenia Z. Ong, Rose Sekulovich, Brian B. Sullivan, Kelly Lindert, Sean B. Sullivan, Pad Chivukula, Steven G. Hughes, Eng Eong. Ooi

**Affiliations:** Singapore General Hospital; Duke-National University of Singapore Medical School; Nanyang Technological University; Arcturus Therapeutics

## Abstract

**Background:** The pandemic of coronavirus disease-19 (Covid-19) continues to afflict the lives and livelihoods of many as global demand for vaccine supply remains unmet.

**Methods:** Phase 1 of this trial (N=42) assessed the safety, tolerability and immunogenicity of ascending levels of one-dose ARCT-021, a self-amplifying mRNA vaccine against Covid-19. Phase 2 (N=64) tested two-doses of ARCT-021 given 28 days apart. Both young and older adults were enrolled. The primary safety outcomes were local and systemic solicited adverse events (AEs) reported immediately and up to 7 days post-inoculation and unsolicited events reported up to 56 days after inoculation. Secondary and exploratory outcomes were antibody and T cell responses to vaccination, respectively.

**Results:** ARCT-021 was well tolerated up to one 7.5 μg dose and two 5.0 μg doses. Local solicited AEs, namely injection-site pain and tenderness, as well as systemic solicited AEs, such as fatigue, headache and myalgia, were more common in ARCT-021 than placebo recipients, and in younger than older adults. Seroconversion rate for anti-S IgG was 100% in all cohorts except for the 1 μg one-dose in younger adults and the 7.5 μg one-dose in older adults, which were each 80%. Neutralizing antibody titers increased with increasing dose although the responses following 5.0 μg and 7.5 μg ARCT-021 were similar. Anti-S IgG titers overlapped with those in Covid-19 convalescent plasma. ARCT-021 also elicited T-cell responses against the S glycoprotein.

**Conclusion:** Taken collectively, the favorable safety and immunogenicity profiles support further clinical development of ARCT-021.

## Introduction

The coronavirus disease 2019 (Covid-19) pandemic, caused by the severe acute respiratory syndrome coronavirus-2 (SARS-CoV-2), has devastated lives globally^1^. Accelerated vaccine deployment has started to reverse this trend^2,3^. The most effective vaccines have been mRNA vaccines^4-6^. A newer synthetic mRNA platform - self-amplifying RNA (sa-mRNA) - may have an added advantage^7,8^; *in situ* mRNA amplification may allow for smaller doses to be administered, stretching the reach of limited vaccine supply to greater number of people^9^. However, there remains a paucity of data that informs on the safety and immunogenicity of such a vaccine construct.

We conducted a first-in-human clinical trial in healthy adults to evaluate the safety, tolerability and immunogenicity of ARCT-021, a self-transcribing and replicating mRNA (STARR™) vaccine candidate for the prevention of Covid-19. The mRNA is a replicon that comprises the Venezuelan equine encephalitis virus (VEEV) genome in which the structural genes have been replaced with the SARS-CoV-2 full-length spike (S) gene, and formulated with the proprietary LUNAR^®^ lipid nanoparticle (LNP). Translation of the replicon produces a multi-protein replicase complex that amplifies a subgenomic mRNA for elevated expression of the S glycoprotein. Preclinical experiments showed that a single dose of ARCT-021 elicited strong Th1-predominant humoral and cellular immune responses against the S protein that protected K-18 human ACE2 transgenic mice from lethal SARS-CoV-2 challenge^10^. Here we report the findings from the Phase 1/2 clinical trial.

## Methods

### Trial design, participants and oversight

ARCT-021-01 is a randomized, double-blinded, placebo (0.9% saline) controlled study to assess the safety, tolerability and immunogenicity of different dose levels of ARCT-021. The primary endpoint was safety and tolerability; secondary and exploratory endpoints were antibody and T-cell responses. The trial was conducted at the Singapore Health Services (SingHealth) Investigational Medicine Unit, following approvals by the SingHealth Centralized Institutional Review Board (CIRB F/2020/2553) and the Singapore Health Sciences Authority. The trial was registered in clinicaltrial.gov (NCT04480957). A safety review committee reviewed data regarding safety and overall trial progress, including dose escalation decisions.

The trial comprised two overlapping parts and evaluated a range of doses of ARCT-021 versus placebo given as one- or two-dose administration to younger (21 to 55 years) and older (56 to 80 years) adult participants. In the Phase 1 part, a one-dose administration (dose levels 1.0 μg, 5.0 μg, 7.5 μg and 10 μg ARCT-021 versus placebo) was given as an intramuscular (IM) injection to younger adults (Cohorts A, B, D1 and C, respectively) and a single dose (7.5 μg ARCT-021 versus placebo) was given to older adults (Cohort D2). In the Phase 2 part, two-dose administrations of 3.0 μg and 5.0 μg ARCT-021 versus placebo separated by 28 days were administered to younger participants (Cohorts F and E) and older participants (Cohorts H and G), respectively. All participants were followed up for 56 days after the last study vaccine administration.

Full lists of the inclusion and exclusion criteria are provided in the protocol. Written informed consent was provided by all the participants before enrollment.

Arcturus Therapeutics, Inc. was the regulatory sponsor of the trial. Both Arcturus Therapeutics and Duke-NUS Medical School co-designed the clinical trial and were responsible for the collection, analysis, and interpretation of the data and for the writing of the report. Arcturus Therapeutics and the corresponding author had full access to all the data in the trial and had final responsibility for the decision to submit the manuscript for publication. All the trial data were available to all the authors. Clinical monitoring, pharmacovigilance, and data management were performed by the Contract Research Organization, CTI.

### Trial procedures

Participants were randomized after completing all screening assessments and eligibility criteria. Phase 1 participants were randomized 5:2 to receive ARCT-021 or placebo. Phase 2 participants were randomized 3:1 to receive ARCT-021 or placebo. All participants were administered 0.5-ml injections of ARCT-021 or placebo into the lateral aspect of the deltoid muscle of the non-dominant arm while the second injection (Phase 2 cohorts) was administered into the contralateral arm. All participants were observed for a minimum of 4 hours after the injection. Blood samples were obtained for safety and immunogenicity assessments according to protocol schedule. The initial planned single doses to be tested were 1.0 μg, 5.0 μg, 10 μg and 20 μg in the single dose cohorts; The final tested doses were 1.0 μg, 5.0 μg, 7.5 μg and 10 μg in the single dose cohorts and 3.0ug and 5.0ug in the two dose cohorts.

### Vaccine and placebo

ARCT-021 was presented as a sterile, frozen, aqueous formulation with 0.2 mg/mL of mRNA-2002 and as a 1.0 mL fill (0.2 mg/1 mL) in 2-mL Type I glass vials, stored frozen at -70°C (+/- 10°C). It is a white to off-white liquid when thawed with a nominal pH of 8.0 and osmolality of approximately1300 mOsm/kg. The placebo was 0.9% sterile saline provided by the study center.

### Safety Assessments

Local and systemic solicited and unsolicited AEs were recorded daily by the participant in a symptom diary for at least 7 days and up to 14 days post vaccination if any events persisted beyond Day 7. The symptom diaries were reviewed by site staff at study visits for up to 14 days post each injection. Injection site was inspected at all visits up to Day 15 (for first injection)/Day 43 (for second injection) or until resolution of local reactogenic event(s). Unsolicited events were collected at all visits and for the duration of study participation.

### Immunogenicity

IgM, IgA and IgG against full-length recombinant S protein were assessed using a Luminex immuno-assay. IgG against the S protein subdomains (RBD, NTD and S2) were also measured for 5.0μg, 7.5μg single and 5.0μg expansion cohorts. Neutralizing antibody was measured using plaque reduction neutralization test (PRNT), with a clinical SARS CoV-2 isolate (hCoV-19/Singapore/2/2020), and the serum titer that neutralizes 50% of the virus inoculum (PRNT_50_) was calculated as previously described^11^. SARS-CoV-2 specific T-cell responses were assessed using flow cytometry (FC) and IFNγ ELISPOT assay following stimulation with overlapping S protein peptide pools.

### Statistical Analysis

No formal sample size calculation was performed. Based on experience from previous studies with other RNA based therapies, the chosen cohort sizes were considered sufficient to meet the objectives of the study while minimizing unnecessary participant exposure. For analysis of safety and humoral immunogenicity, placebo participants were pooled by age group and number of doses administered as follows: A to D1 pooled, E and F pooled, G and H pooled. D2 was the only single dose older adult cohort so D2 placebos were not pooled. For humoral immunogenicity, confidence intervals of the geometric means were calculated with the Student’s t distribution on log-transformed data. Seroconversion was defined as at least a 4-fold increase in antibody titer from baseline.

## Results

### Trial Participants

In total, 169 healthy volunteers were screened, with 106 participants randomized and injected. The distribution of participants by cohort is shown in Figure 1. All participants received assigned doses of ARCT-021. All enrolled participants completed the planned study scheduled trial visits. A summary of demographic characteristics by treatment assignment is presented in Table 1.

**Figure 1.**
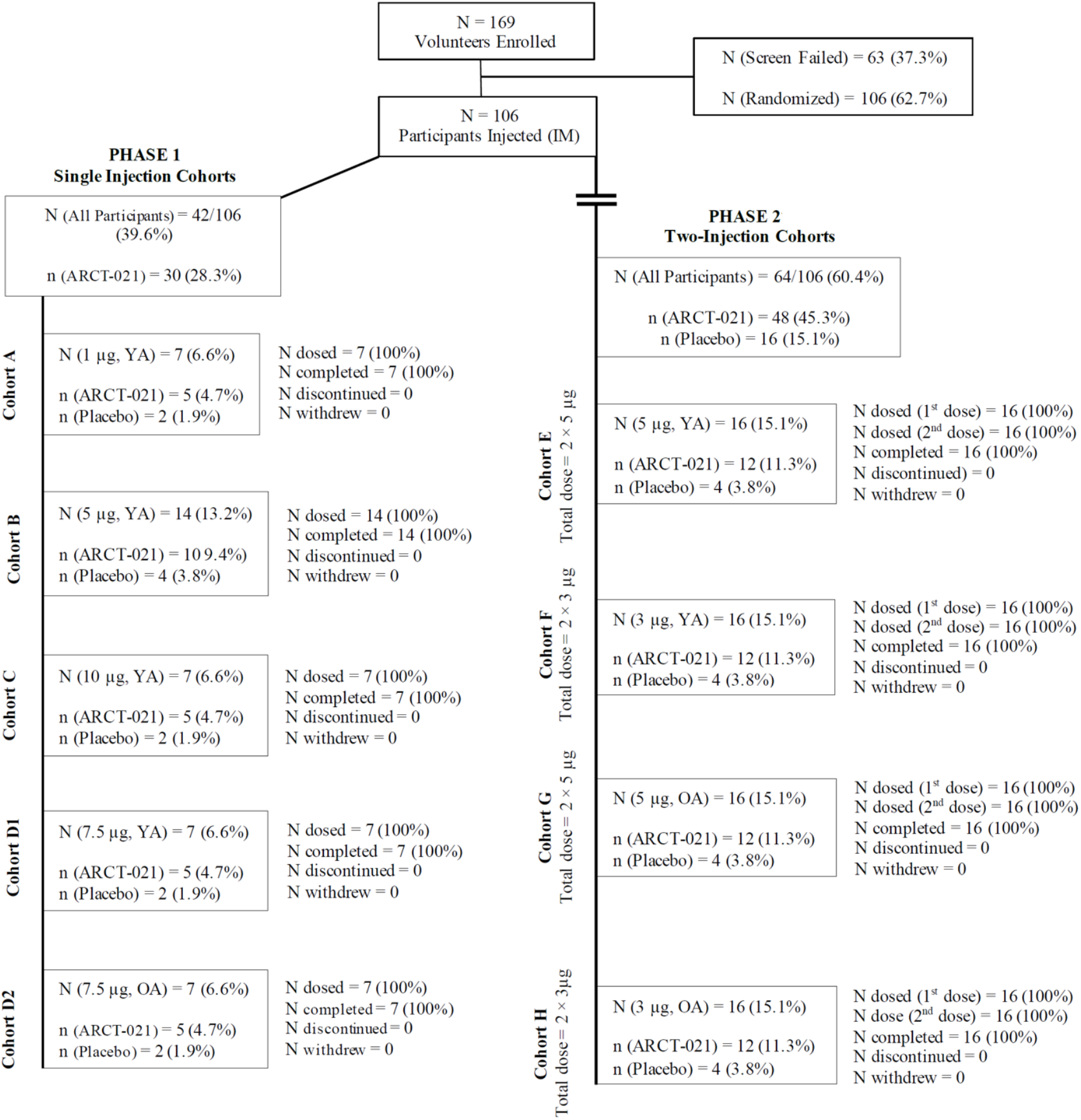
Enrolment and randomization of trial participants. Abbreviations: N = number of participants in the cohort; n = number of participants in each treatment group; OA = older adults (56-80 years); YA = younger adults (21-55 years). Phase 1 – Single injection, escalating dose. Study duration 56 days post first injection. Phase 2 – Two same dose injections, 28 days apart. Study duration 85 days post first injection.

**Table 1.** Demographic Characteristics of the Participants, According to Dose Cohort and Age Group*.

| Cohort | Single Dose Cohorts |  |  |  |  |  |  | Two-Dose Cohorts |  |  |  |  |  |
| --- | --- | --- | --- | --- | --- | --- | --- | --- | --- | --- | --- | --- | --- |
|  | Age 21 - 55 |  |  |  |  | Age 56 - 80 |  | Age 21 - 55 |  |  | Age 56 - 80 |  |  |
| Variable | Cohort<br>A<br>1µg | Cohort<br>B<br>5µg | Cohort<br>C<br>10µg | Cohort<br>D1<br>7.5µg | Pooled<br>Placebo<br>Cohorts<br>A to D1 | Cohort<br>D2<br>7.5µg | Cohort<br>D2<br>Placebo | Cohort<br>E<br>5µg | Cohort<br>F<br>3µg | Pooled<br>Placebo<br>Cohorts<br>E and F | Cohort<br>G<br>5µg | Cohort<br>H<br>3µg | Pooled<br>Placebo<br>Cohorts<br>G and H |
| Number of Participants | 5 | 10 | 5 | 5 | 10 | 5 | 2 | 12 | 12 | 8 | 12 | 12 | 8 |
| Sex - n (%) |  |  |  |  |  |  |  |  |  |  |  |  |  |
| Male | 3<br>(60.0) | 6<br>(60.0) | 2<br>(40.0) | 4<br>(80.0) | 8<br>(20.0) | 5<br>(100) | 1<br>(50.0) | 8<br>(66.7) | 10<br>(83.3) | 5<br>(62.5) | 11<br>(91.7) | 8<br>(66.7) | 7<br>(87.5) |
| Female | 2<br>(40.0) | 4<br>(40.0) | 3<br>(60.0) | 1<br>(20.0) | 2<br>(20.0) | 0<br>(0) | 1<br>(50.0) | 4<br>(33.3) | 2<br>(16.7) | 3<br>(37.5) | 1<br>(8.3) | 4<br>(33.3) | 1<br>(12.5) |
| Race - n (%)† |  |  |  |  |  |  |  |  |  |  |  |  |  |
| Asian | 5<br>(100) | 10<br>(100) | 5<br>(100) | 4<br>(80.0) | 10<br>(100) | 5<br>(100) | 2<br>(100) | 11<br>(91.7) | 10<br>(83.3) | 7<br>(87.5) | 12<br>(100) | 12<br>(100) | 7<br>(87.5) |
| White | 0<br>(0) | 0<br>(0) | 0<br>(0) | 1<br>(20.0) | 0<br>(0) | 0<br>(0) | 0<br>(0) | 0<br>(0) | 1<br>(8.3) | 1<br>(12.5) | 0<br>(0) | 0<br>(0) | 1<br>(12.5) |
| American Indian or Alaskan<br>Native | 0<br>(0) | 0<br>(0) | 0<br>(0) | 0<br>(0) | 0<br>(0) | 0<br>(0) | 0<br>(0) | 1<br>(8.3) | 0<br>(0) | 0<br>(0) | 0<br>(0) | 0<br>(0) | 0<br>(0) |
| Other | 0<br>(0) | 0<br>(0) | 0<br>(0) | 0<br>(0) | 0<br>(0) | 0<br>(0) | 0<br>(0) | 0<br>(0) | 1<br>(8.3) | 0<br>(0) | 0<br>(0) | 0<br>(0) | 0<br>(0) |
| Hispanic Ethnic Group – n (%)† | 0<br>(0) | 0<br>(0) | 0<br>(0) | 0<br>(0) | 0<br>(0) | 0<br>(0) | 0<br>(0) | 0<br>(0) | 0<br>(0) | 0<br>(0) | 0<br>(0) | 0<br>(0) | 0<br>(0) |
| Age (years) ‡ |  |  |  |  |  |  |  |  |  |  |  |  |  |
| Mean (SD) | 35<br>(±9.17) | 39.5<br>(±7.15) | 41.4<br>(±12.48) | 40.4<br>(±10.41) | 33.5<br>(±8.72) | 64<br>(±5.61) | 61.5<br>(±6.36) | 34.5<br>(±8.33) | 38<br>(±9.34) | 39.1<br>(±7.95) | 62.3<br>(±4.63) | 61.7<br>(±3.77) | 63.3<br>(±5.70) |
|  | Age 21 - 55 |  |  |  |  | Age 56 - 80 |  | Age 21 - 55 |  |  | Age 56 - 80 |  |  |
| Variable | Cohort<br>A<br>1µg | Cohort<br>B<br>5µg | Cohort<br>C<br>10µg | Cohort<br>D1<br>7.5µg | Pooled<br>Placebo<br>Cohorts<br>A to D1 | Cohort<br>D2<br>7.5µg | Cohort<br>D2<br>Placebo | Cohort<br>E<br>5µg | Cohort<br>F<br>3µg | Pooled<br>Placebo<br>Cohorts<br>E and F | Cohort<br>G<br>5µg | Cohort<br>H<br>3µg | Pooled<br>Placebo<br>Cohorts<br>G and H |
| Median [range] | 32.0<br>[28-50] | 40.0<br>[29-51] | 44.0<br>[24-55] | 47.0<br>[24-48] | 34.5<br>[21-46] | 64.0<br>[57-70] | 61.5<br>[57-66] | 35.5<br>[21-47] | 35.0<br>[22-51] | 37.0<br>[30-51] | 62.0<br>[56-70] | 61.5<br>[56-68] | 64.5<br>[56-71] |

Abbreviations: N = total number of subjects in each dose cohort; n = number of subjects contributing to the summary; SD = standard deviation
\*Percentages may not total 100 because of rounding
†Race and ethnic group were reported by the participant.
‡ The age of the participants was the age at the time of screening

### Safety

A summary of solicited local and systemic adverse events (AE) is presented in Figures 2 and 3. ARCT-021 was generally well tolerated up to the 7.5 μg dose. The 10 μg dose was associated with more local and systemic solicited AE, including grade 3 severity. Although the study stopping rules were not met, the protocol was amended to remove further dose escalation. A younger (Cohort D1) and older adult (Cohort D2) one-dose cohorts at the 7.5 μg dose level were added following safety review. A single serious adverse event due to cellulitis of the foot in a placebo recipient following an insect-bite was reported. It was medically attended and resolved after two days. All participants in the 2-dose cohorts completed the vaccinations without any delay.

**Figure 2.**
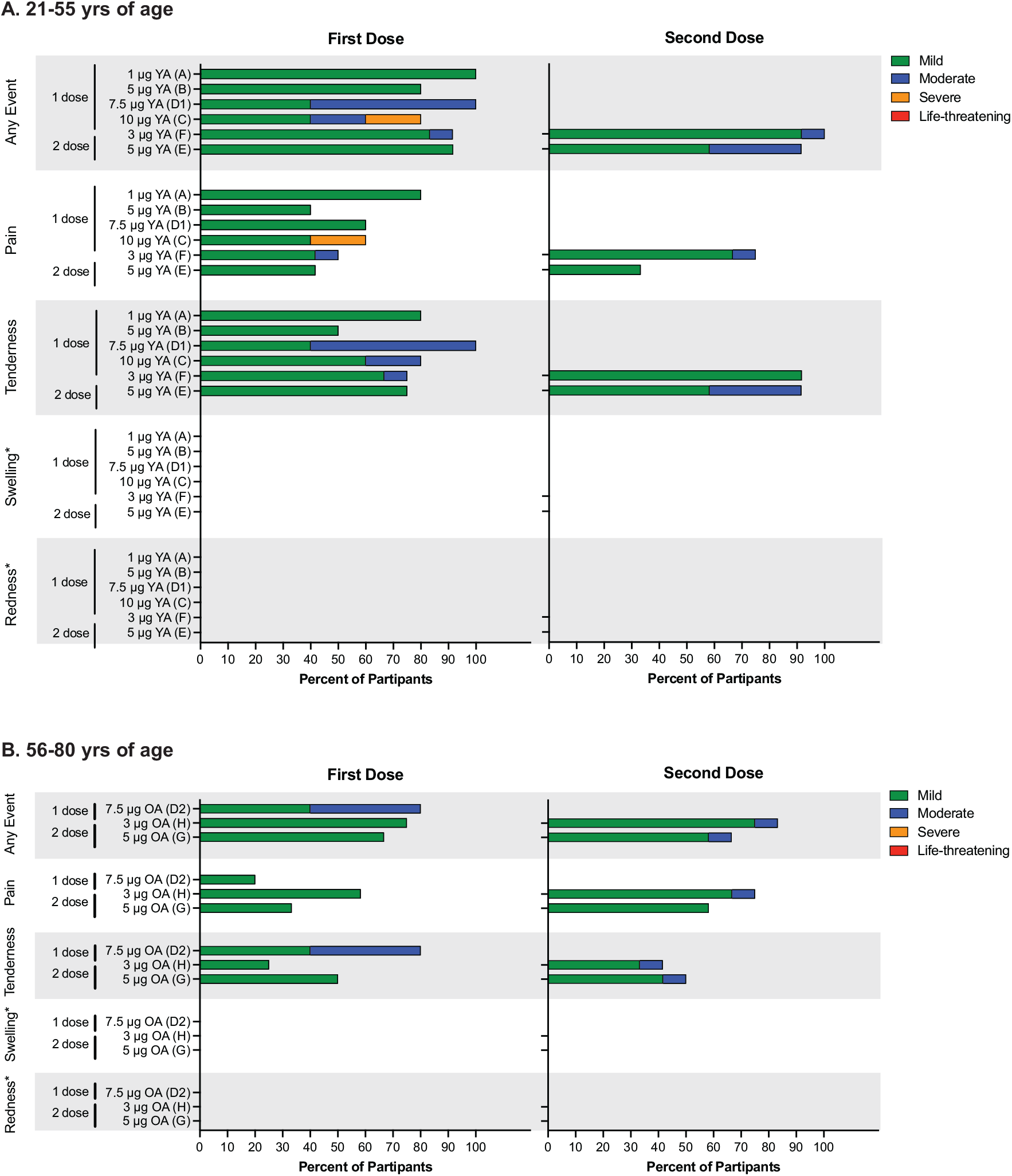
Local solicited adverse events. Shown are the percentages of vaccinated participants reporting local solicited events. Data are represented for age groups, 21-55 years (Panel A) and 56-80 years (Panel B). Severity of local solicited Events are displayed as mild (green), moderate (blue), severe (orange) or life-threatening (red). *Note, injection site swelling and injection site redness were not reported. YA=Younger Adults (21-55); OA=Older Adults (56-80).

**Figure 3.**
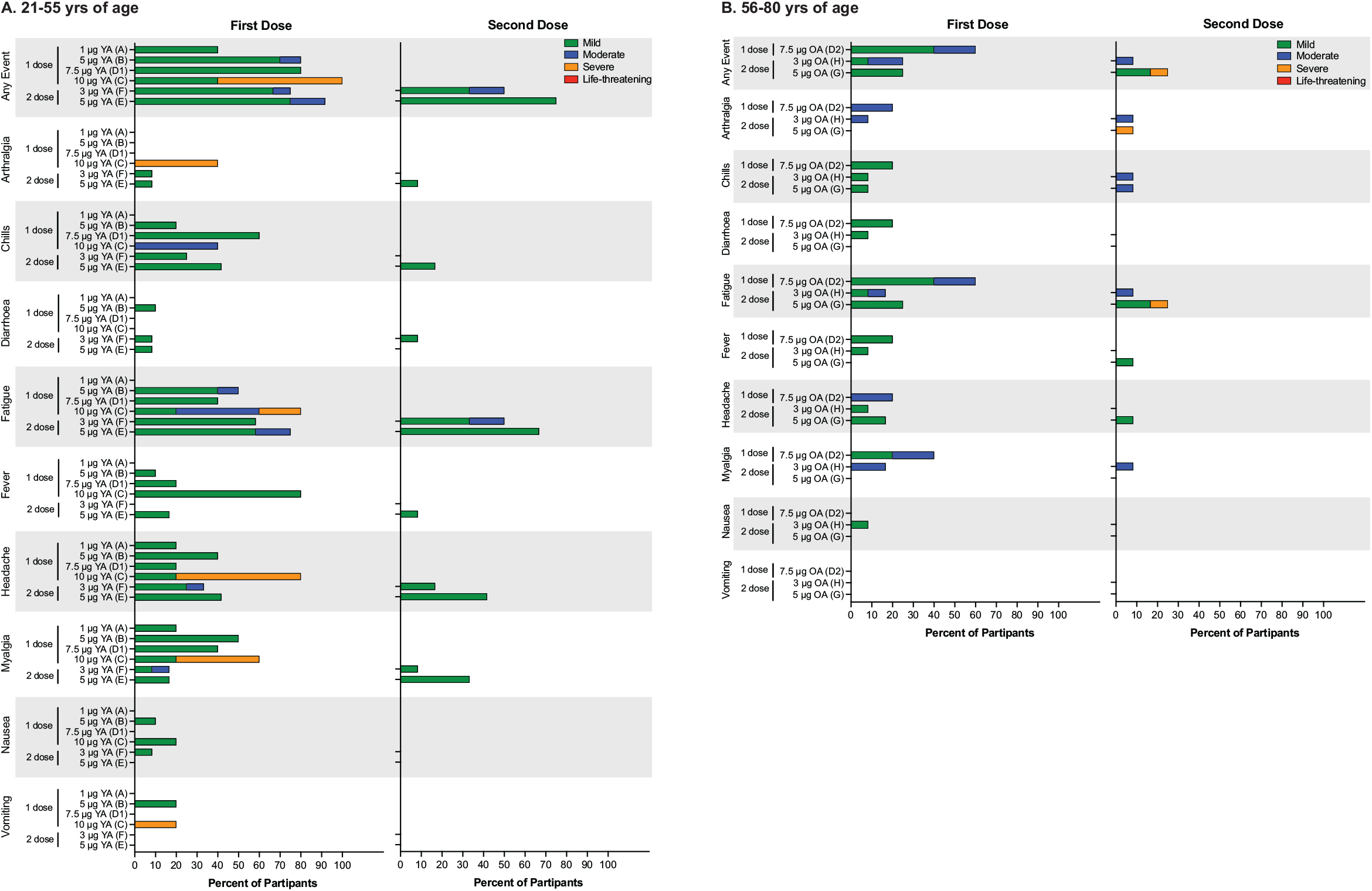
Systemic solicited adverse events. Percentage of systemic solicited events reported in vaccinated participants aged 21-55 years (Panel A) and 56-80 years (Panel B). Severity of systemic solicited Events are displayed as mild (green), moderate (blue), severe (orange) or life-threatening (red). YA=Younger Adults (21-55); OA=Older Adults (56-80).

### Solicited Treatment Emergent Adverse Events

#### Local reactions

Sixty-nine (88.5%) participants who received ARCT-021 reported local solicited AEs, as compared to 21.4% of placebo-treated participants (Figure 2). These included injection site tenderness and pain in 73.1% and 60.3% of participants, respectively. In the two-dose younger adult cohorts, local solicited AE occurred in 11(91.7%) following the first dose at both 5.0 μg and 3.0 μg and in 11 (91.7%) and 12 (100%), respectively, following the second dose. In older adults, local solicited events occurred in 8 (66.7%) and 9 (75%) following the first dose and in 8 (66.7%) and 10 (83.3%) following the second dose in the 5.0 μg and 3.0 μg dose cohorts, respectively. A higher proportion of participants experienced moderate grade injection site tenderness with the second dose. At the same dose level, a higher proportion of younger than older adults experienced ≥ grade 2 solicited AE. There was no reported injection site erythema or swelling. There was one grade 3 injection site pain at the 10μg dose. There were no grade 4 local reactions and no discernable dose relationship.

#### Systemic Reactions

Overall, 62.8% of participants who received ARCT-021 reported a systemic solicited AE as compared to 46.4% amongst the placebo recipients (Figure 3). The most common events were fatigue (50.0%), headache (34.6%), myalgia (28.2%), chills (25.6%), and fever (14.1%) and were more common in younger than older adults. Systemic AEs were primarily mild or moderate with no grade 4 events reported. Grade 3 solicited events occurred primarily in the 10 μg dose cohort. Two grade 3 events (fatigue and arthralgia) occurred in a single participant following the second dose in the 5.0 μg older adult cohort. Severity of systemic AEs trended with dose. Most systemic AEs commenced within 1 to 2 days after vaccination and resolved or reduced in severity within 7 days.

### Unsolicited Systemic Treatment Emergent Adverse Events

Unsolicited AEs are summarized in Supplementary Table 1. In general, more participants vaccinated with ARCT-021 reported unsolicited AEs after vaccination. Five were severe, occurring primarily in the 10 μg dose cohort. These events occurred on the day after injection and resolved or improved in severity within 2 days after onset. One participant in older adult 3.0 μg two dose cohort (F) experienced grade 3 transaminitis 57 days after the last dose of ARCT-021. This event was classed as unlikely related by the Investigator and resolved after 18 days. No severe events occurred in placebo treated participants.

### Abnormal hematological and biochemical findings

There appeared to be a dose-related trend for ≥grade 2 lymphopenia with 0%, 25%, 26.5%, 30.0% and 40.0% of participants affected at the 1.0 μg, 3.0 μg, 5.0 μg, 7.5 μg and 10 μg dose levels, respectively. Onset of lymphopenia occurred within 24 hours after injection and resolved uneventfully, generally within a day. The incidence was similar following the first (14 of 78 participants [17.9%]) and second dose (10 of 48 participants [20.8%]). Three of 48 (6.3%) participants had ≥ grade 2 lymphopenia following both the first and second injection. There was no trend for increased severity following the second injection. The incidence was similar in older (24.1%) and younger adults (26.5%). There were 10 (12.8%) Grade 2 neutropenia events, the incidence of which was higher in older (17.2%) than younger (10.2%) adults. All neutropenic episodes were asymptomatic, resolved spontaneously and did not result in any sequelae.

Post-baseline elevations of ALT occurred in 5 ARCT-021 treated participants (Supplementary Table 3). Two of these participants had ALT elevation at screening, which had normalized at Day 1. No elevations of hepatic enzymes were observed in placebo participants. There were no other notable laboratory abnormalities.

### Immunogenicity

#### SARS-CoV-2 Binding Antibodies and Neutralization Response

Seroconversion rate for anti-S IgG was 100% in all cohorts except for the one-dose younger adult cohort A (1μg) and the one-dose older adult cohort D2 (7.5 μg), which were both 80%. The IgG response increased with increasing dose (Fig 4A, Table S5 and S6), although there was a trend for better responses at the 5.0 μg dose than at the 7.5 μg dose in older adults after a single dose (compared at Day 29 in the two-dose cohorts). With the exception of the 1 μg dose, S-binding IgG titers overlapped with the range of antibodies in Covid-19 convalescent plasma (Fig. 4A). Vaccine-induced IgG antibodies bound the various S protein subdomains, including the RBD, NTD and the highly conserved S2 (Fig. 4B). PRNT_50_ titers increased with increasing dose, while the responses were similar following one-dose of 5.0 μg and 7.5 μg. At doses ≥3 μg, PRNT_50_ titers were within the range observed in convalescent sera from patients with mild to moderate Covid-19^10^.

**Figure 4.**
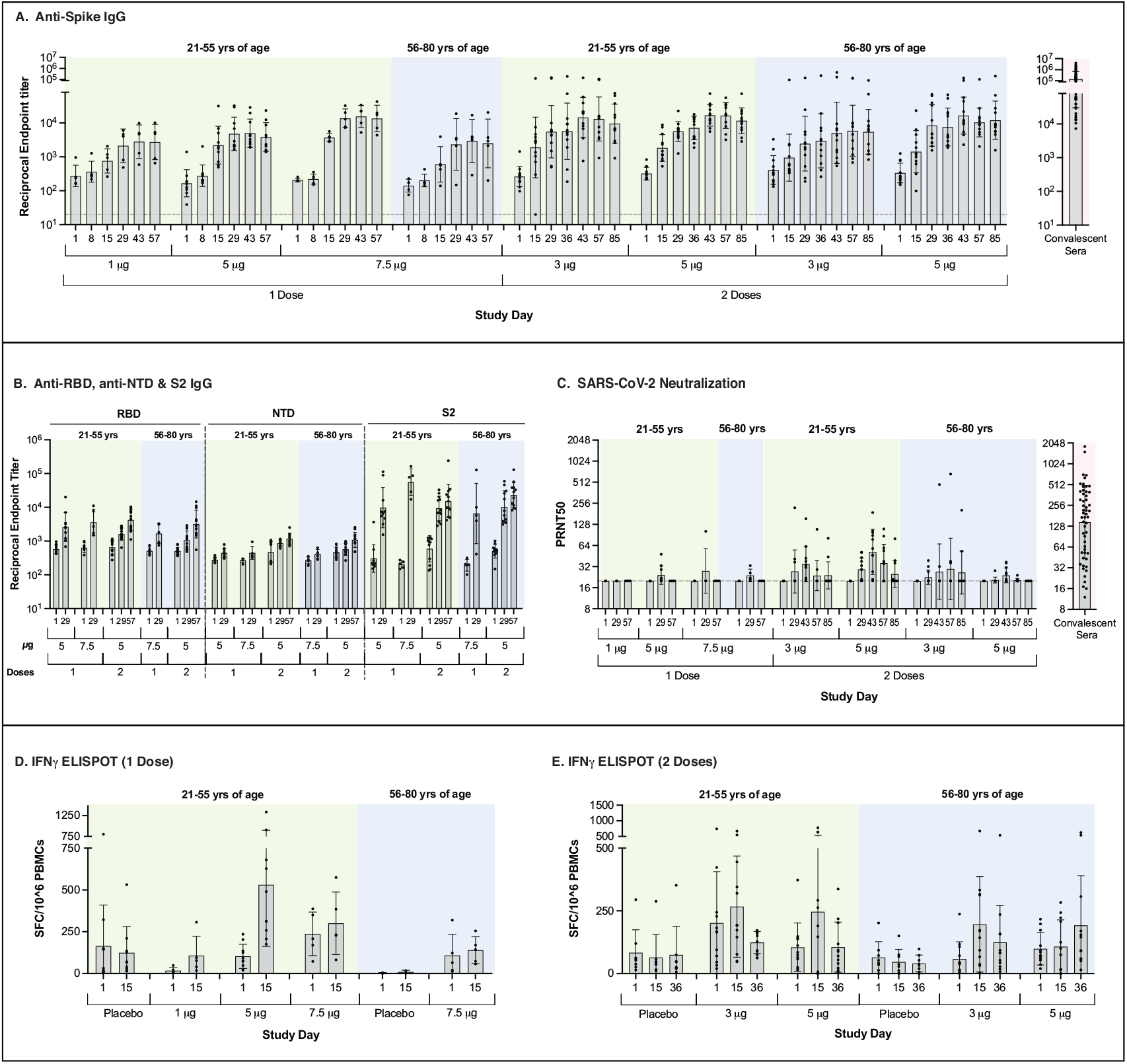
Antibody and T cell responses. Shown are IgG reciprocal endpoint titers to SARS-CoV-2 whole ectodomain Spike using Luminex immune-Assay (Panel A). IgG reciprocal titers to the Spike sub-domains, i.e. receptor-binding domain (RBD), N-terminal binding domain (NTD) and domain 2 (S2) using Luminex immuno-assay is represented for selected cohorts and timepoints (Panel B). Antibody neutralization of live SARS-CoV-2 virus was measured using a BSL3 plaque reduction neutralization assay (PRNT) and represented as 50% neutralization titers (PRNT50) (Panel C). Cellular immunity and T cell responses in prime and prime-boost cohorts were measured using an interferon gamma (IFNγ) enzyme-linked immune absorbent spot (ELISPOT) assay and represented as spot-forming cells (SFC) per million PBMCs (Panel D and E). Displayed ELISPOT results are cumulative responses of six peptide pools spanning the entire SARS-CoV-2 Spike. Peptide pool-specific responses presented in Fig. S2 and S3. For Panels A-C, the error bars display standard deviation around the GMT depicted by a horizontal bar. For Panels D and E, the whiskers display standard deviation (SD) around the mean (horizontal bar). Each dot represents an individual participant.

#### SARS-CoV-2 T-Cell Response

T-cell responses were observed on both ELISPOT and ICS in response to stimulation with six peptide pools covering the entire S glycoprotein (Fig. S1). IFNγ ELISPOT responses were generally maximal at Day 15, and were similar at the 5.0 μg and 7.5 μg dose in younger adults (Fig. 4D-E and Fig. S2). Responses in the 5.0 μg older adult cohort peaked after the second dose (Fig. 4E and Fig. S3). IFNγ ELISPOT responses were generally higher in younger adults (Fig. 4D-E). CD4 T-cell IFN-γ responses as measured by ICS showed the greatest values at Day 29 in the 1.0 μg and 5.0 μg single dose cohorts (Fig. S5). CD4 T-cell IFN-γ responses were generally greater in older adults than younger adults at the 5.0 μg and 7.5 μg doses and were similar between the two age groups at the 3 μg dose. A further increase in CD4 T-Cell IFN-γ responses following second dose was only seen in the 5.0 μg younger adults. At all tested doses, the CD4 T-cell responses were Th1 dominant (Fig. S6). The CD8 T-cell IFN-γ cytokine responses by ICS showed the highest values at Day 29 in the 5.0 μg single dose cohort. CD8 IFN-γ responses in the older adult cohorts were higher than the responses observed in the younger adult cohorts at the same dose (Fig. S4). There was no further increase in CD8 T-cell IFN-γ responses following the second dose (Fig. S4).

## Discussion

The shortage in Covid-19 vaccine supply continues to affect global recovery from this pandemic. A dose-sparing sa-mRNA vaccine could thus contribute to addressing this shortage.

We found an overall favorable safety profile up to 7.5ug dose. Over 96% of all AEs were either mild or moderate. Transient lymphopenia and neutropenia, possibly due to innate immune driven redistribution of lymphocytes^12^ were observed, similar to other mRNA vaccines^13,14^.

ARCT-021 is also immunogenic. Anti-S IgG antibody levels were within the range of convalescent plasma, even after just one-dose. Vaccination also produced anti-S antibody of different types – IgM and IgA – in addition to IgG. These antibodies can elicit Fc-mediated functions, such as complement activation, antibody-dependent cellular cytotoxicity (ADCC) and phagocytosis, as well as protect the mucosa. Indeed, recent reports suggest that Covid-19 can be prevented despite low levels^15^ or even absence of neutralizing antibodies^16^. Even antibodies that enhance SARS-CoV-2 infection *in vitro* protected non-human primates (NHPs) against SARS-CoV-2 challenge^17^. Finally, onset of mRNA vaccine-mediated protection against Covid-19 was associated with the development of anti-S antibodies that showed ADCC activity, along with S-reactive T cells, without neutralizing antibodies^11,18^. These findings collectively indicate that other functions of antibodies besides virus neutralization play key roles in protecting against Covid-19.

Although a single dose of ARCT-021 was immunogenic, the second dose did not boost antibody titers dramatically. This may be due to three reasons: Firstly, the one-month interval between prime and boost may be too short for maximal boosting; secondly, we have not modified the S gene based on the immunogenicity and efficacy observed in our preclinical study^10^ and; finally, we wanted to ensure that our CD8 T cell response would be directed against the native S since, at the time when ARCT-021 development began, the CD8 T cell epitopes had not been fully mapped.

Our intention to elicit appreciable CD8 T cell responses with vaccination was driven by the collective body of evidence indicating the importance of cellular immunity in preventing Covid-19. Pre-clinical studies on ARCT-021 showed that vaccine-induced protection against SARS-CoV-2 was mediated primarily by CD8 T-cells rather than B cells^10^. In NHPs, cellular immunity compensated for low antibody levels in protecting against SARS-CoV-2 infection^19^. Clinically, delayed T but not B cell responses were associated with severe disease^20^. Recognition of epitopes spanning the S protein by CD8 T cells could also protect against neutralizing antibody-evading variants of concern (VOC)^21^; several such VOCs are presently spreading globally and will be the main challenge for any new Covid-19 vaccines^22^. Indeed, T cell responses may underpin the sustained efficacy of mRNA vaccines against these VOCs^23^.

Our findings contrast with the phase 1 trial finding of another sa-mRNA vaccine^24^. The reasons for the different outcomes are uncertain currently although there are differences in both the S gene sequence and vaccine formulation^10,25^. Further investigations to understand the underpinnings of different immunogenicity outcome could be informative on future sa-mRNA vaccine design.

Taken collectively, the favorable safety and immunogenicity profiles support further clinical development of ARCT-021. A larger multi-center phase 2 clinical trial (NCT04668339) recruiting ∼600 healthy adult volunteers is ongoing to guide a subsequent phase 3 clinical trial.

## Data Availability

Data available upon request and with necessary approvals from the corresponding author.

## Supplementary Appendix

**Table S1:** Summary of Unsolicited Treatment Emergent Adverse Events According to Dose Cohort and Age Group.

| Cohort | Single Dose |  |  |  |  |  |  | Two-Dose |  |  |  |  |  |
| --- | --- | --- | --- | --- | --- | --- | --- | --- | --- | --- | --- | --- | --- |
|  | Age 21 - 55 |  |  |  |  | Age 56 - 80 |  | Age 21 - 55 |  |  | Age 56 - 80 |  |  |
| Event Category / n(%) | Cohort A<br>1µg<br>(N=5) | Cohort B<br>5µg<br>(N=10) | Cohort C<br>10µg<br>(N=5) | Cohort D1<br>7.5µg<br>(N=5) | Pooled<br>Placebo<br>Cohorts<br>A to D1<br>(N=10) | Cohort D2<br>7.5µg<br>(N=5) | Cohort D2<br>Placebo<br>(N=2) | Cohort E<br>5µg<br>(N=12) | Cohort F<br>3µg<br>(N=12) | Pooled<br>Placebo<br>Cohorts<br>E and F<br>(N=8) | Cohort G<br>5µg<br>(N=12) | Cohort H<br>3µg<br>(N=12) | Pooled<br>Placebo<br>Cohorts<br>G and H<br>(N=8) |
| Any Unsolicited Event | 4 (80.0) | 7 (70.0) | 5 (100.0) | 4 (80.0) | 5 (50.0) | 5 (100.0) | 0 | 8 (66.7) | 8 (66.7) | 6 (75.0) | 12 (100.0) | 8 (66.7) | 3 (37.5) |
| Related | 4 (80.0) | 6 (60.0) | 5 (100.0) | 3 (60.0) | 0 | 5 (100.0) | 0 | 7 (58.3%) | 8 (66.7%) | 1 (12.5%) | 9 (75.0%) | 5 (41.7%) | 1 (12.5%) |
| Severe | 0 | 0 | 2 (40.0) | 0 | 0 | 0 | 0 | 0 | 0 | 0 | 0 | 1 (8.3) | 0 |
| Life threatening | 0 | 0 | 0 | 0 | 0 | 0 | 0 | 0 | 0 | 0 | 0 | 0 | 0 |
| Any Serious Unsolicited Event | 0 | 0 | 0 | 0 | 1 (10.0) | 0 | 0 | 0 | 0 | 0 | 0 | 0 | 0 |
| Related | 0 | 0 | 0 | 0 | 0 | 0 | 0 | 0 | 0 | 0 | 0 | 0 | 0 |
| Severe | 0 | 0 | 0 | 0 | 0 | 0 | 0 | 0 | 0 | 0 | 0 | 0 | 0 |
| Life-threatening | 0 | 0 | 0 | 0 | 0 | 0 | 0 | 0 | 0 | 0 | 0 | 0 | 0 |
| Any Unsolicited Event Leading to<br>Withdrawal | 0 | 0 | 0 | 0 | 0 | 0 | 0 | 0 | 0 | 0 | 0 | 0 | 0 |
| Related | 0 | 0 | 0 | 0 | 0 | 0 | 0 | 0 | 0 | 0 | 0 | 0 | 0 |
| Severe | 0 | 0 | 0 | 0 | 0 | 0 | 0 | 0 | 0 | 0 | 0 | 0 | 0 |
| Life-threatening | 0 | 0 | 0 | 0 | 0 | 0 | 0 | 0 | 0 | 0 | 0 | 0 | 0 |
| Death | 0 | 0 | 0 | 0 | 0 | 0 | 0 | 0 | 0 | 0 | 0 | 0 | 0 |

**Table S2:**
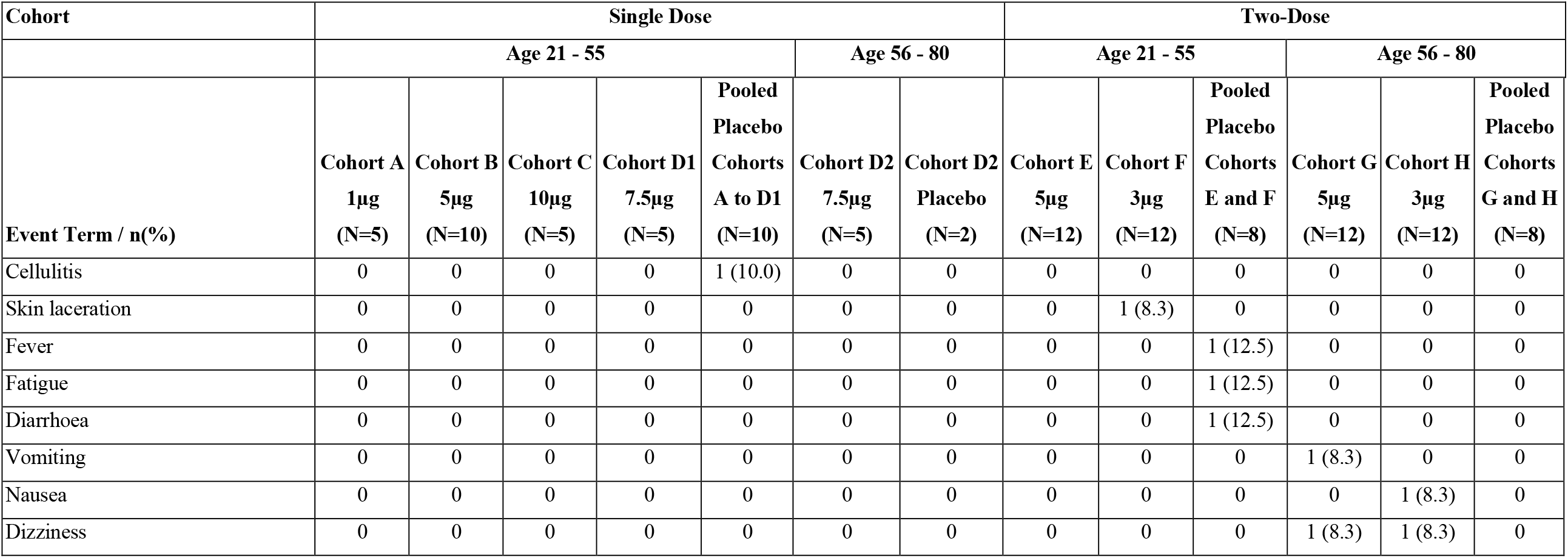
Medically Attended Events by Dose Cohort and Age Group.

**Table S3:**
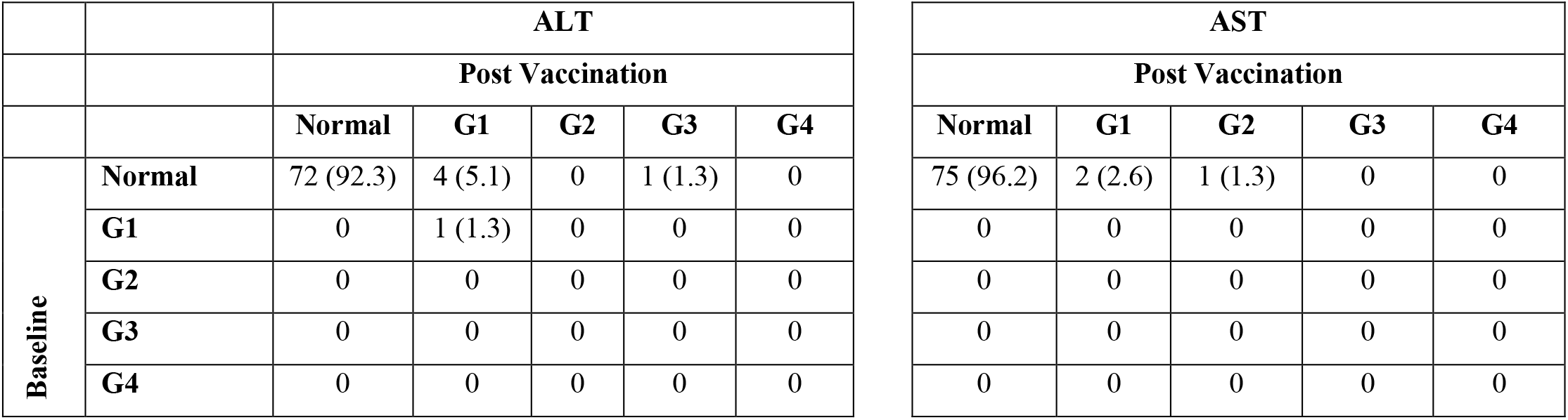
Grade Shifts in Alanine Transaminase and Aspartate Transaminase in ARCT-021 Treated Participants.

|  |  | ALT |  |  |  |  |
| --- | --- | --- | --- | --- | --- | --- |
|  |  | Post Vaccination |  |  |  |  |
|  |  | Normal | G1 | G2 | G3 | G4 |
| Baseline | Normal | 72 (92.3) | 4 (5.1) | 0 | 1 (1.3) | 0 |
|  | G1 | 0 | 1 (1.3) | 0 | 0 | 0 |
|  | G2 | 0 | 0 | 0 | 0 | 0 |
|  | G3 | 0 | 0 | 0 | 0 | 0 |
|  | G4 | 0 | 0 | 0 | 0 | 0 |

| AST |  |  |  |  |
| --- | --- | --- | --- | --- |
| Post Vaccination |  |  |  |  |
| Normal | G1 | G2 | G3 | G4 |
| 75 (96.2) | 2 (2.6) | 1 (1.3) | 0 | 0 |
| 0 | 0 | 0 | 0 | 0 |
| 0 | 0 | 0 | 0 | 0 |
| 0 | 0 | 0 | 0 | 0 |
| 0 | 0 | 0 | 0 | 0 |

**Table S4:**
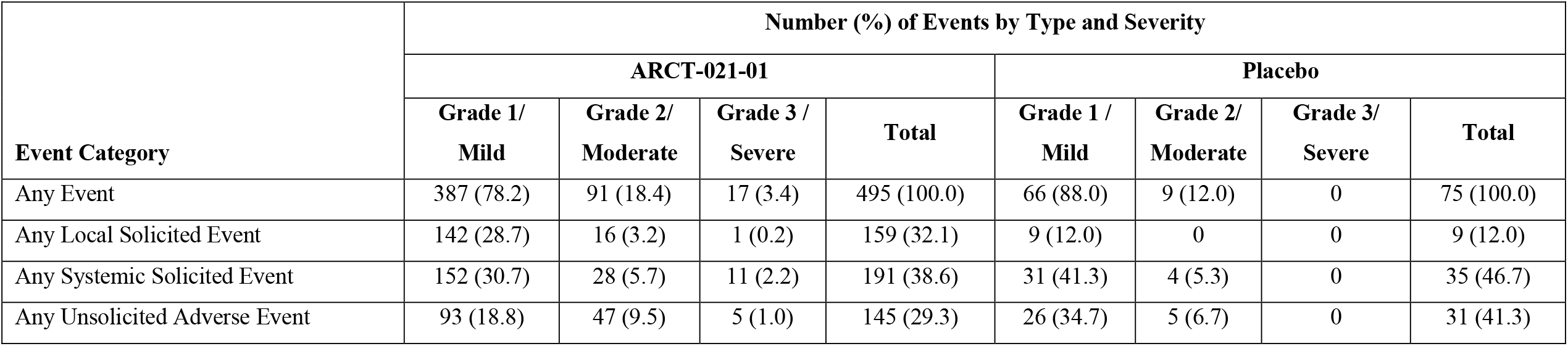
Total Number (%) of Events by Class of Event, Treatment Group and Severity – Safety Population.

**Table S5.** Antibody responses in 1-Dose Cohorts.

|  | Age 21 – 55 |  |  |  |  |  |  |  |  |  | Age 56 – 80 |  |  |  |
| --- | --- | --- | --- | --- | --- | --- | --- | --- | --- | --- | --- | --- | --- | --- |
|  | **1 µg**<br>**(Cohort A)**<br>**N=5** |  | **5 µg**<br>**(Cohort B)**<br>**N=10** |  | **7.5 µg**<br>**(Cohort D1)**<br>**N=5** |  | **10.0 µg**<br>**(Cohort C)**<br>**N=5** |  | **Pooled Placebo**<br>**(Cohorts A, B,**<br>**C, D1)**<br>**N=10** |  | **7.5 µg**<br>**(Cohort D2)**<br>**N=5** |  | **Cohort D2**<br>**Placebo**<br>**N=2** |  |
| **Test/Time point** | **n** | **GMT**<br>**(95% CI)** | **No** | **GMT**<br>**(95% CI)** | **n** | **GMT**<br>**(95% CI)** | **n** | **GMT**<br>**(95% CI)** | **n** | **GMT**<br>**(95% CI)** | **n** | **GMT**<br>**(95% CI)** | **n** | **GMT**<br>**(95% CI)** |
| **Luminex anti-S**<br>**IgG** |  |  |  |  |  |  |  |  |  |  |  |  | 2 |  |
| Day 1 | 5 | 274.0<br>(111.06,<br>676.22) | 10 | 166.3<br>(86.17,<br>321.01) | 5 | 207.5<br>(178.89,<br>240.80) | 5 | 511.7<br>(61.69,<br>4244.31) | 10 | 223.7<br>(126.60,<br>395.45) | 5 | 141.0<br>(81.90,<br>242.68) | 2 | 287.0<br>(NA) |
| Day 8 | 5 | 364.9<br>(151.09,<br>881.32) | 10 | 277.1<br>(163.72,<br>468.84) | 5 | 221.0<br>(145.26,<br>336.15) | 5 | 995.4<br>(54.55,<br>18164.73) | 10 | 278.1<br>(171.81,<br>450.11) | 5 | 201.6<br>(117.94,<br>344.69) | 2 | 398.1<br>(NA) |
| Day 15 | 5 | 757.8<br>(271.89,<br>2112.02) | 10 | 2217.8<br>(888.14,<br>5538.12) | 5 | 3698.4<br>(2628.51,<br>5203.68) | 5 | 9482.5<br>(787.96,<br>114114.7) | 10 | 267.1<br>(162.13,<br>440.09) | 5 | 601.4<br>(135.44,<br>2670.21) | 2 | 408.2<br>(NA) |
| Day 29 | 5 | 2130.0<br>(516.21,<br>8788.98) | 10 | 4781.1<br>(2121.61,<br>10774.20) | 5 | 13675.7<br>(6209.51,<br>30119.08) | 5 | 32748.6<br>(4963.84,<br>216057.2) | 10 | 236.5<br>(133.33,<br>419.49) | 5 | 2333.3<br>(265.53,<br>20502.83) | 2 | 392.7<br>(NA) |
|  | 1 µg<br>(Cohort A)<br>N=5 |  | 5 µg<br>(Cohort B)<br>N=10 |  | 7.5 µg<br>(Cohort D1)<br>N=5 |  | 10.0 µg<br>(Cohort C)<br>N=5 |  | Pooled Placebo<br>(Cohorts A, B,<br>C, D1)<br>N=10 |  | 7.5 µg<br>(Cohort D2)<br>N=5 |  | Cohort D2<br>Placebo<br>N=2 |  |
| Test/Time point | n | GMT<br>(95% CI) | No | GMT<br>(95% CI) | n | GMT<br>(95% CI) | n | GMT<br>(95% CI) | n | GMT<br>(95% CI) | n | GMT<br>(95% CI) | n | GMT<br>(95% CI) |
| Day 36 | 5 | 2542.6<br>(585.27,<br>11046.15) | 10 | 4189.3<br>(2212.27,<br>7933.24) | 5 | 14898.7<br>(5117.38,<br>43375.93) | 5 | 26403.5<br>(5567.92,<br>125207.5) | 10 | 289.4<br>(175.29,<br>477.68) | 5 | 2119.5<br>(252.79,<br>17770.67) | 2 | 330.5<br>(NA) |
| Day 43 | 5 | 2804.2<br>(680.03,<br>11563.45) | 10 | 4958.6<br>(2484.11,<br>9898.07) | 5 | 15480.5<br>(6111.05,<br>39215.12) | 5 | 28763.3<br>(4965.70,<br>166608.3) | 10 | 298.2<br>(171.99,<br>517.04) | 5 | 2942.3<br>(477.76,<br>18120.22) | 2 | 528.8<br>(NA) |
| Day 56 | 5 | 2734.4<br>(616.36,<br>12130.49) | 10 | 3843.1<br>(1912.88,<br>7721.23) | 5 | 13573.3<br>(4462.61,<br>41284.14) | 5 | 21768.1<br>(3916.06,<br>121002.1) | 10 | 297.2<br>(151.31,<br>583.74) | 5 | 2490.5<br>(318.10,<br>19498.39) | 2 | 415.0<br>(NA) |
| **Live Virus**<br>**PRNT<sub>50</sub>** |  |  |  |  |  |  |  |  |  |  |  |  |  |  |
| Day 1 | 5 | 20.0<br>(20.00,<br>20.00) | 10 | 20.0<br>(20.00,<br>20.00) | 5 | 20.0<br>(20.00,<br>20.00) | 5 | 20.0<br>(20.00,<br>20.00) | 10 | 20.0<br>(20.00,<br>20.00) | 5 | 20.0<br>(20.00,<br>20.00) | 2 | 20.0<br>(NA) |
|  | 1 µg<br>(Cohort A)<br>N=5 |  | 5 µg<br>(Cohort B)<br>N=10 |  | 7.5 µg<br>(Cohort D1)<br>N=5 |  | 10.0 µg<br>(Cohort C)<br>N=5 |  | Pooled Placebo<br>(Cohorts A, B,<br>C, D1)<br>N=10 |  | 7.5 µg<br>(Cohort D2)<br>N=5 |  | Cohort D2<br>Placebo<br>N=2 |  |
| Test/Time point | n | GMT<br>(95% CI) | No | GMT<br>(95% CI) | n | GMT<br>(95% CI) | n | GMT<br>(95% CI) | n | GMT<br>(95% CI) | n | GMT<br>(95% CI) | n | GMT<br>(95% CI) |
| Day 29 | 5 | 20.0<br>(20.00,<br>20.00) | 10 | 24.2<br>(19.50,<br>29.95) | 5 | 27.7<br>(11.21,<br>68.46) | 5 | 32.9<br>(14.73,<br>73.56) | 10 | 20.9<br>(19.19,<br>22.68) | 5 | 24.0<br>(18.61,<br>30.89) | 2 | 20.0<br>(NA) |
| Day 57 | 5 | 20.0<br>(20.00,<br>20.00) | 10 | 20.0<br>(20.00,<br>20.00) | 5 | 20.0<br>(20.00,<br>20.00) | 5 | 20.9<br>(18.48,<br>23.67) | 10 | 20.0<br>(20.00,<br>20.00) | 5 | 20.0<br>(20.00,<br>20.00) | 2 | 20.0<br>(NA) |
| **Luminex anti-S**<br>**IgM** |  |  |  |  |  |  |  |  |  |  |  |  |  |  |
| Day 1 | 5 | 50.0<br>(50.00,<br>50.00) | 10 | 73.5<br>(50.77,<br>106.51) | 5 | 84.7<br>(44.63,<br>160.91) | 5 | 77.9<br>(34.93,<br>173.82) | 10 | 67.3<br>(47.40,<br>95.49) | 5 | 53.0<br>(45.06,<br>62.37) | 2 | 73.1<br>(NA) |
| Day 8 | 5 | 53.5<br>(44.37,<br>64.47) | 10 | 73.6<br>(53.95,<br>100.50) | 5 | 63.9<br>(42.06,<br>97.20) | 5 | 140.1<br>(11.23,<br>1749.25) | 10 | 63.3<br>(46.52,<br>86.06) | 5 | 56.1<br>(40.74,<br>77.29) | 2 | 116.4<br>(NA) |
|  | 1 µg<br>(Cohort A)<br>N=5 |  | 5 µg<br>(Cohort B)<br>N=10 |  | 7.5 µg<br>(Cohort D1)<br>N=5 |  | 10.0 µg<br>(Cohort C)<br>N=5 |  | Pooled Placebo<br>(Cohorts A, B,<br>C, D1)<br>N=10 |  | 7.5 µg<br>(Cohort D2)<br>N=5 |  | Cohort D2<br>Placebo<br>N=2 |  |
| Test/Time point | n | GMT<br>(95% CI) | No | GMT<br>(95% CI) | n | GMT<br>(95% CI) | n | GMT<br>(95% CI) | n | GMT<br>(95% CI) | n | GMT<br>(95% CI) | n | GMT<br>(95% CI) |
| Day 15 | 5 | 90.6<br>(28.15,<br>291.26) | 10 | 427.0<br>(141.33,<br>1290.32) | 5 | 215.4<br>(104.49,<br>443.91) | 5 | 646.9<br>(99.38,<br>4211.00) | 10 | 60.8<br>(42.94,<br>86.03) | 5 | 171.1<br>(69.58,<br>420.93) | 2 | 136.4<br>(NA) |
| **Luminex anti-S**<br>**IgA** |  |  |  |  |  |  |  |  |  |  |  |  |  |  |
| Day 1 | 5 | 383.1<br>(36.21,<br>4052.75) | 10 | 142.1<br>(56.96,<br>354.28) | 5 | 101.1<br>(52.46,<br>194.97) | 5 | 236.1<br>(73.94,<br>754.10) | 10 | 111.7<br>(86.18,<br>144.75) | 5 | 61.5<br>(44.63,<br>84.76) | 2 | 88.3<br>(NA) |
| Day 29 | 5 | 2119.1<br>(159.82,<br>28096.25) | 10 | 1259.6<br>(336.74,<br>4711.46) | 5 | 525.3<br>(159.82,<br>1726.38) | 5 | 9056.6<br>(1083.57,<br>75696.55) | 10 | 84.9<br>(64.26,<br>112.21) | 5 | 321.7<br>(88.20,<br>1172.99) | 2 | 162.9<br>(NA) |
| **Luminex anti-**<br>**RBD IgG\*** |  |  |  |  |  |  |  |  |  |  |  |  |  |  |
|  | 1 µg<br>(Cohort A)<br>N=5 |  | 5 µg<br>(Cohort B)<br>N=10 |  | 7.5 µg<br>(Cohort D1)<br>N=5 |  | 10.0 µg<br>(Cohort C)<br>N=5 |  | Pooled Placebo<br>(Cohorts A, B,<br>C, D1)<br>N=10 |  | 7.5 µg<br>(Cohort D2)<br>N=5 |  | Cohort D2<br>Placebo<br>N=2 |  |
| Test/Time point | n | GMT<br>(95% CI) | No | GMT<br>(95% CI) | n | GMT<br>(95% CI) | n | GMT<br>(95% CI) | n | GMT<br>(95% CI) | n | GMT<br>(95% CI) | n | GMT<br>(95% CI) |
| Day 1 |  |  | 10 | 574<br>(467;<br>707) | 5 | 631<br>(411;<br>969) |  |  | 6 | 513<br>(340;<br>774) | 5 | 510<br>(369;<br>705) | 2 | 926<br>(8;<br>113760) |
| Day 29 |  |  | 10 | 2618<br>(1299;<br>5276) | 5 | 3578<br>(1171;<br>10932) |  |  | 6 | 653<br>(389;<br>1098) | 5 | 1665<br>(717;<br>3869) | 2 | 1113<br>(52;<br>23697) |
| **Luminex anti-NTD IgG\*** |  |  |  |  |  |  |  |  |  |  |  |  |  |  |
| Day 1 |  |  | 10 | 276<br>(242;<br>314) | 5 | 269<br>(223;<br>326) |  |  | 6 | 308<br>(198;<br>481) | 5 | 269<br>(202;<br>357) | 2 | 347<br>(61;<br>1964) |
| Day 29 |  |  | 10 | 443<br>(359;<br>548) | 5 | 446<br>(260;<br>766) |  |  | 6 | 323<br>(220;<br>474) | 5 | 420<br>(290;<br>608) | 2 | 309<br>(174;<br>549) |
|  | 1 µg<br>(Cohort A)<br>N=5 |  | 5 µg<br>(Cohort B)<br>N=10 |  | 7.5 µg<br>(Cohort D1)<br>N=5 |  | 10.0 µg<br>(Cohort C)<br>N=5 |  | Pooled Placebo<br>(Cohorts A, B,<br>C, D1)<br>N=10 |  | 7.5 µg<br>(Cohort D2)<br>N=5 |  | Cohort D2<br>Placebo<br>N=2 |  |
| Test/Time point | n | GMT<br>(95% CI) | No | GMT<br>(95% CI) | n | GMT<br>(95% CI) | n | GMT<br>(95% CI) | n | GMT<br>(95% CI) | n | GMT<br>(95% CI) | n | GMT<br>(95% CI) |
| Luminex anti-S2<br>IgG* |  |  |  |  |  |  |  |  |  |  |  |  |  |  |
| Day 1 |  |  | 10 | 305<br>(156;<br>596) | 5 | 204<br>(149;<br>279) |  |  | 6 | 390<br>(144;<br>1055) |  | 192<br>(116;<br>317) | 2 | 276<br>(215;<br>356) |
| Day 29 |  |  | 10 | 9703<br>(3631;<br>25928) | 5 | 55676<br>(18159;<br>170702) |  |  | 6 | 380<br>(152;<br>953) |  | 6520<br>(506;<br>83940) | 2 | 270<br>(101;<br>725) |

S = Full-Length SARS-CoV-2 Spike Protein; S2 = S2 domain of SARS-CoV-2 Spike Protein, RBD = receptor binding domain of SARS-CoV-2 Spike Protein; NTD =N-terminal domain of SARS-CoV-2 Spike Protein; PRNT =Plaque Reduction Neutralization Test; GMT=Geometric Mean Titer; CI=Confidence Interval;
N represents the total number of subjects in each cohort. n represents the number of subjects contributing to the summary.
\*IgG titers against RBD, NTD and S2 not assessed for subjects in Cohorts A and C.

**Table S6.**
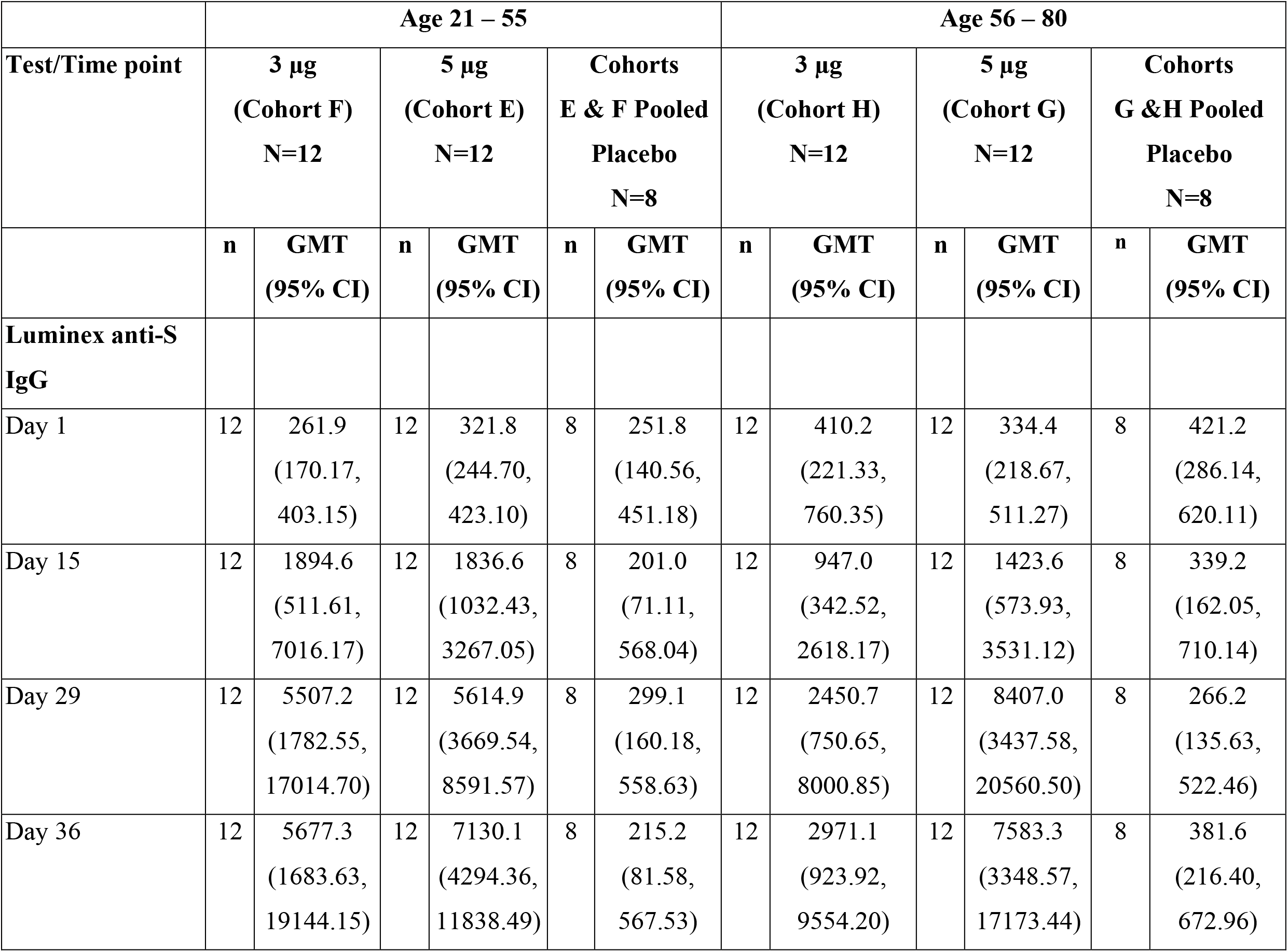
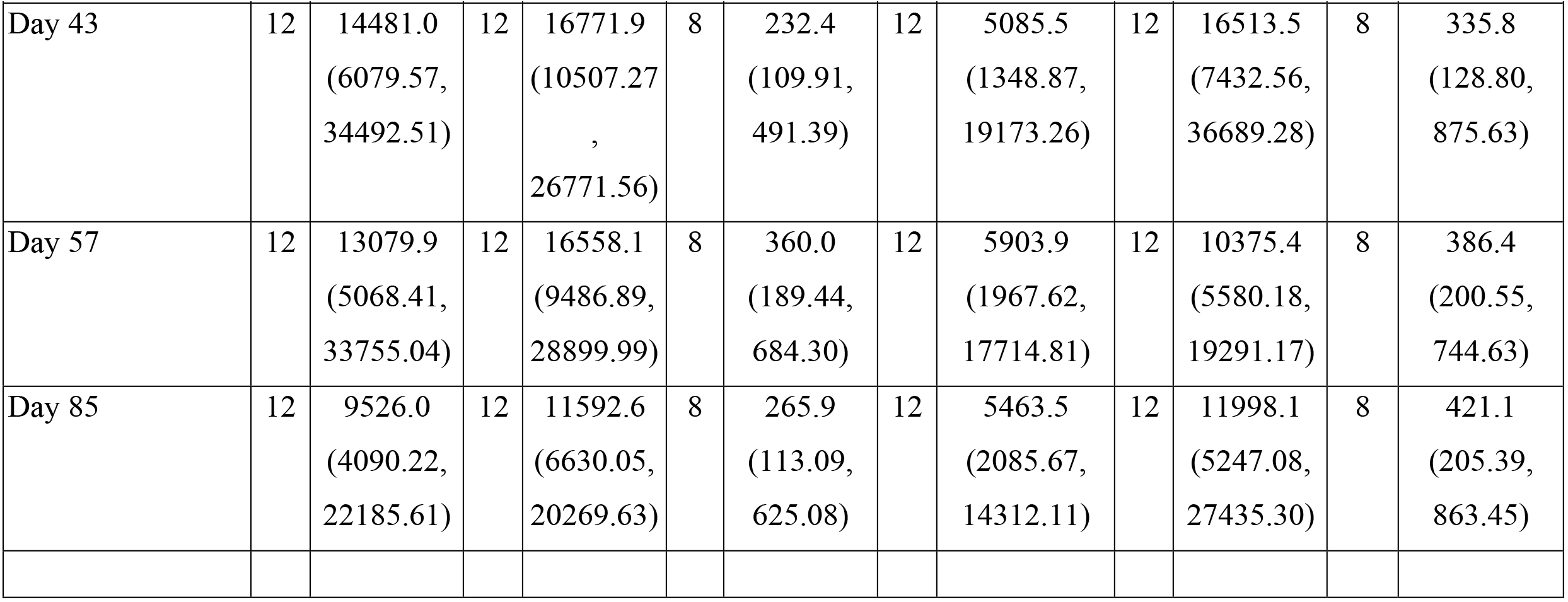
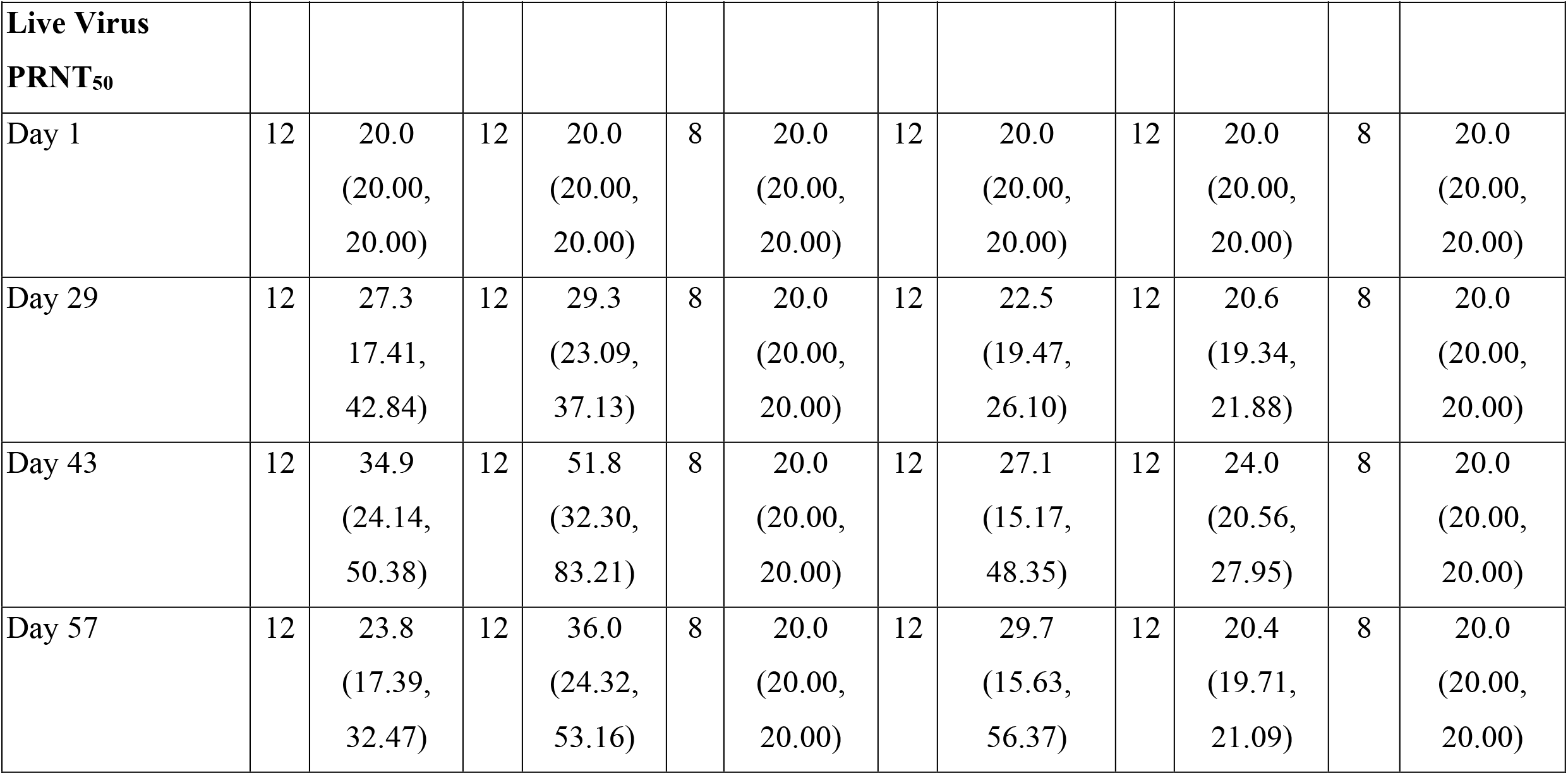
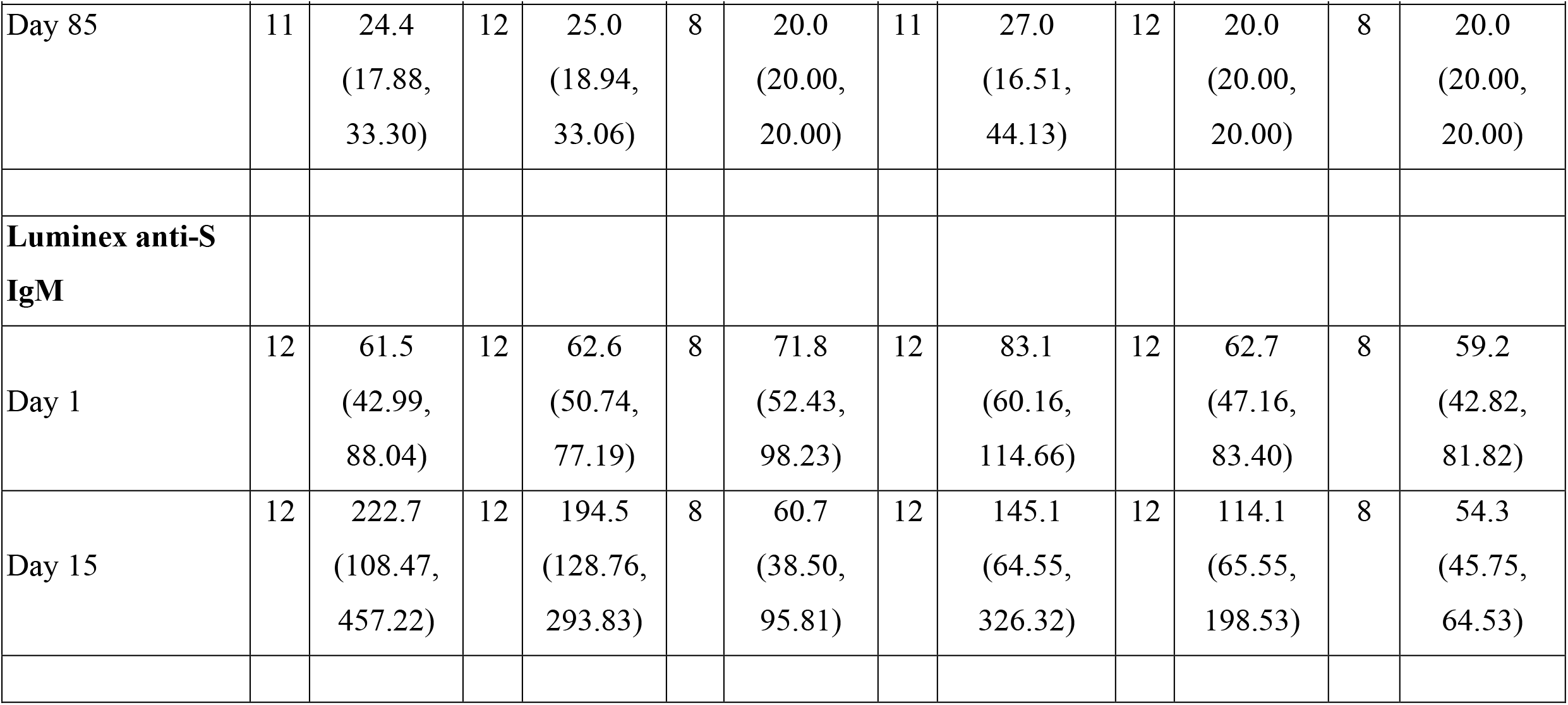
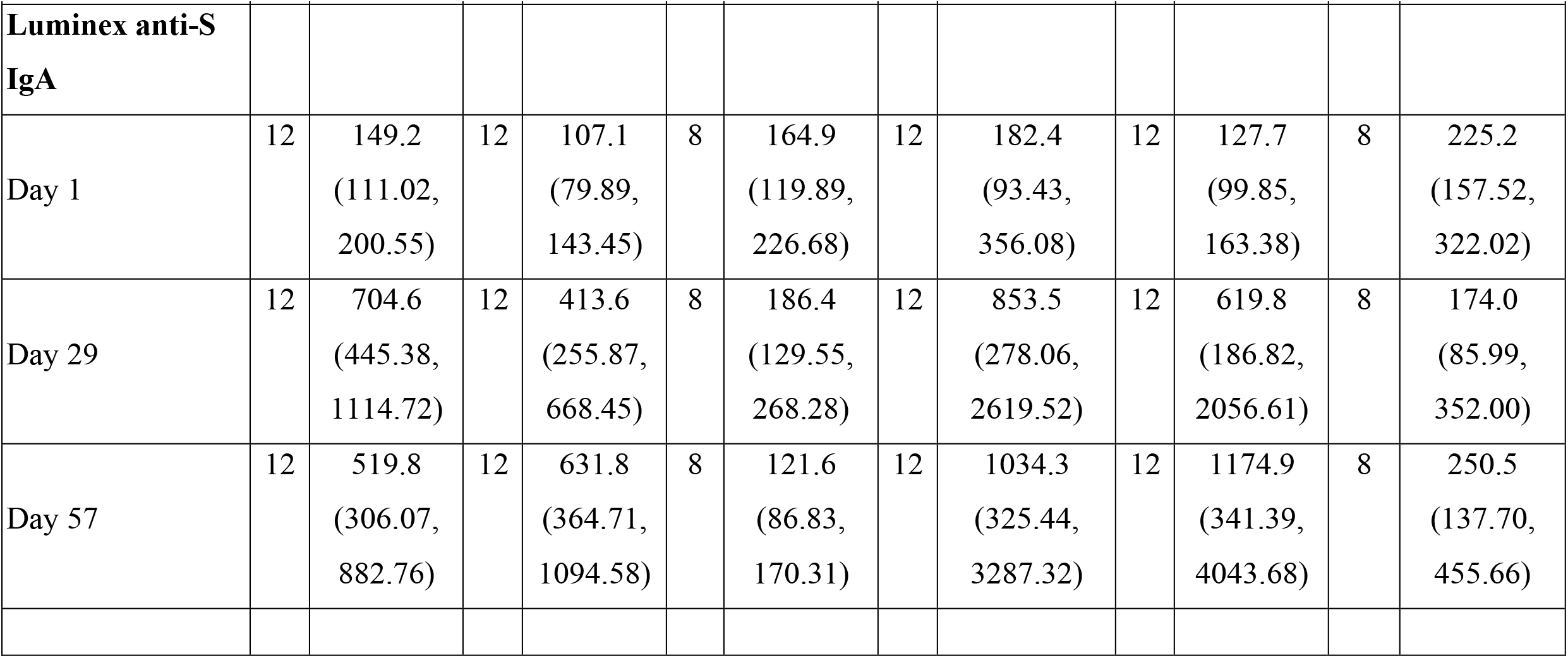
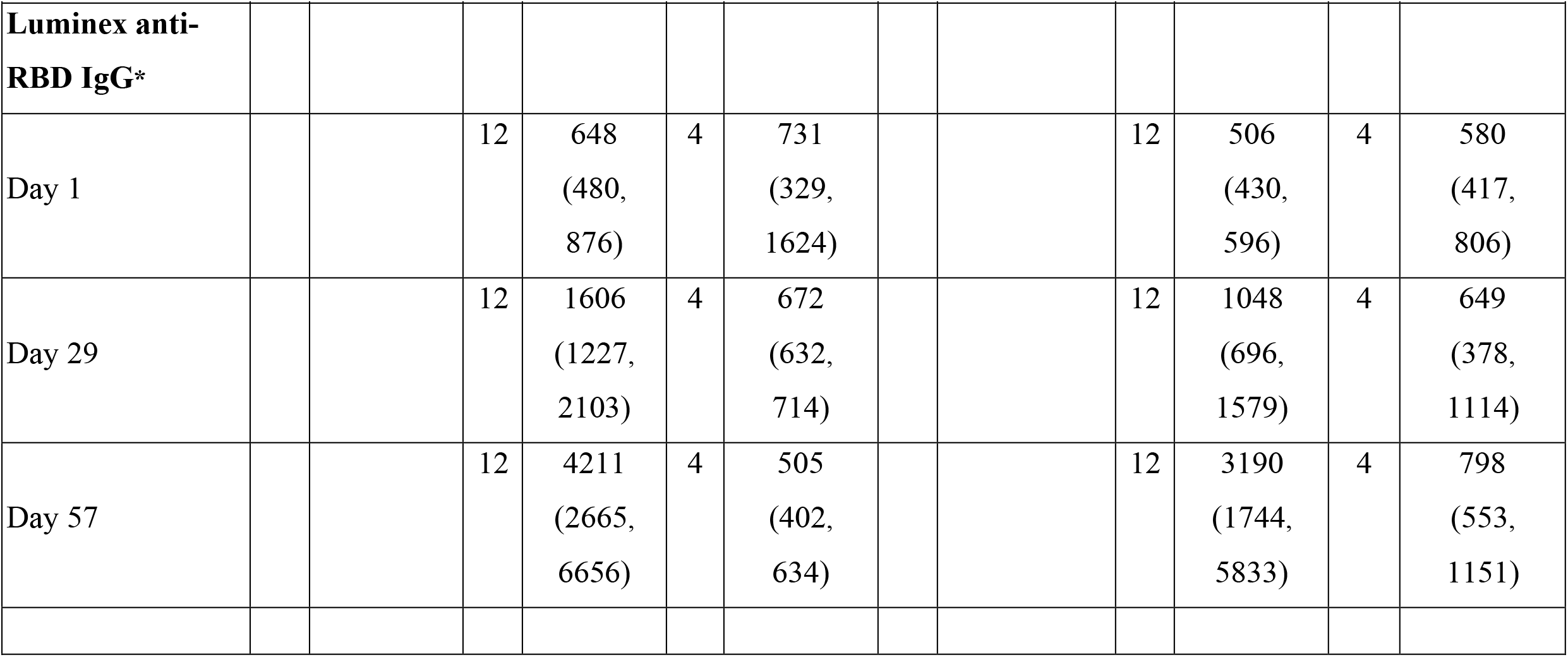
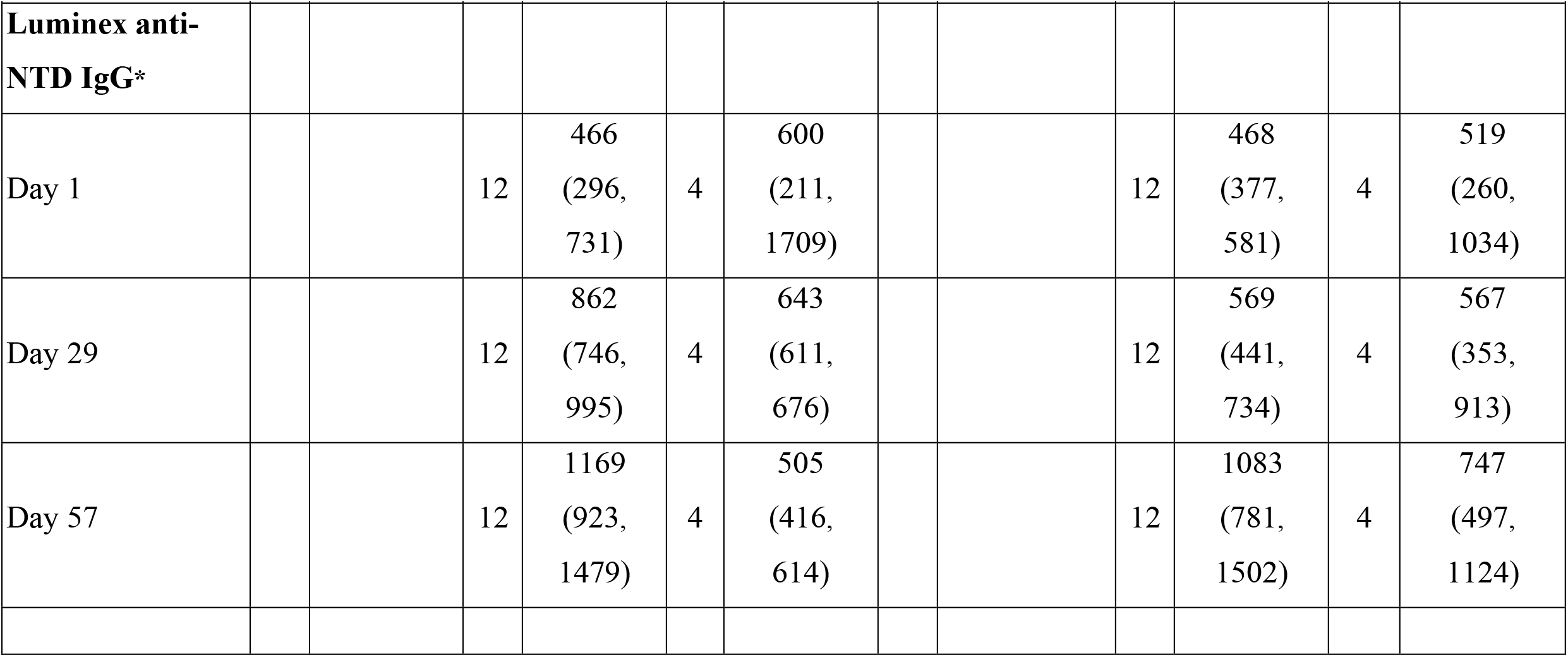
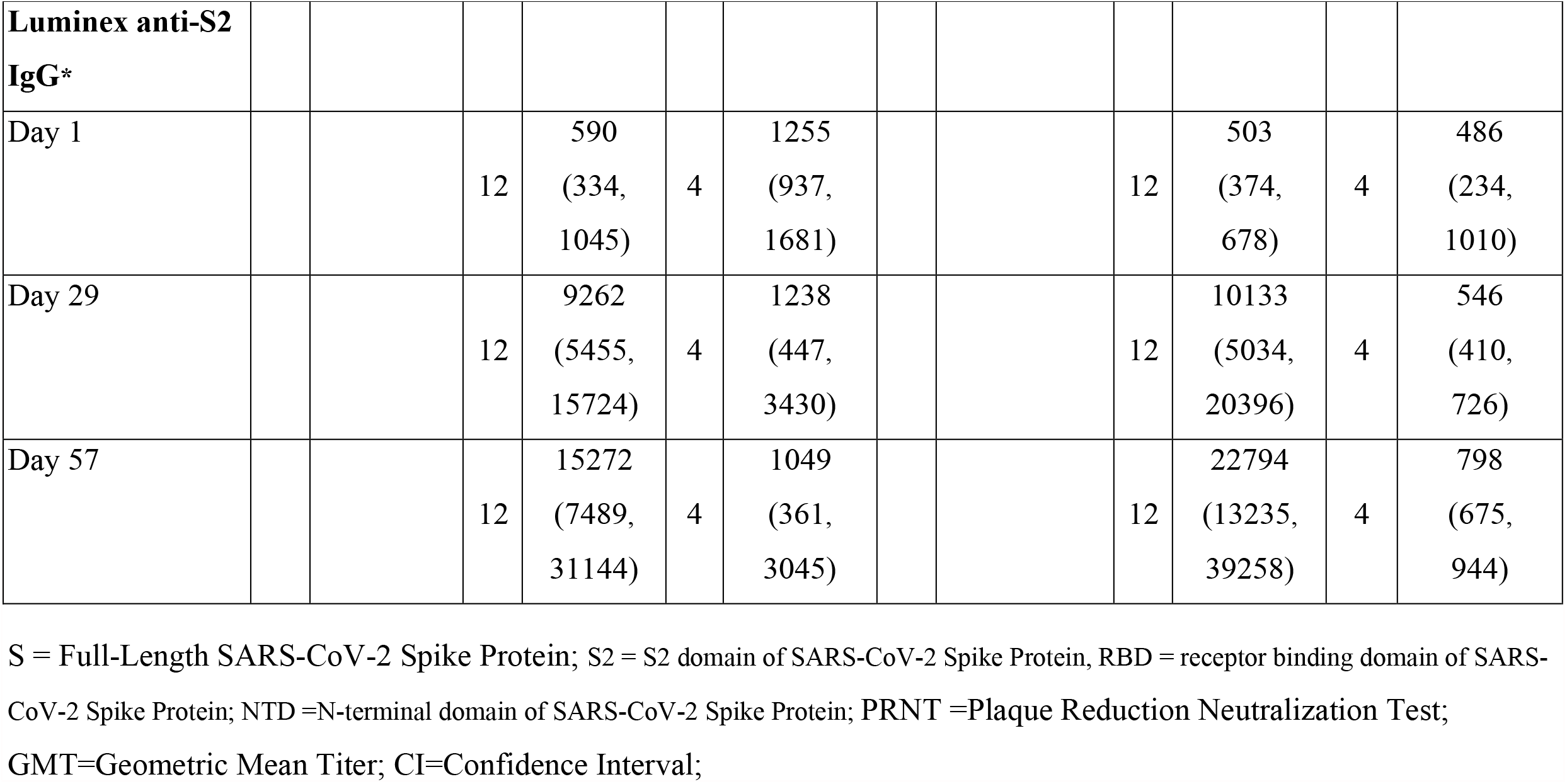
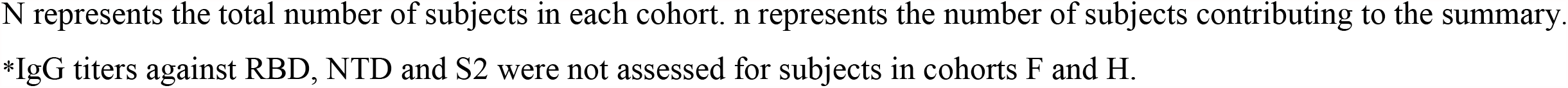
Antibody responses in 2-Dose Cohorts.

**Figure S1.**
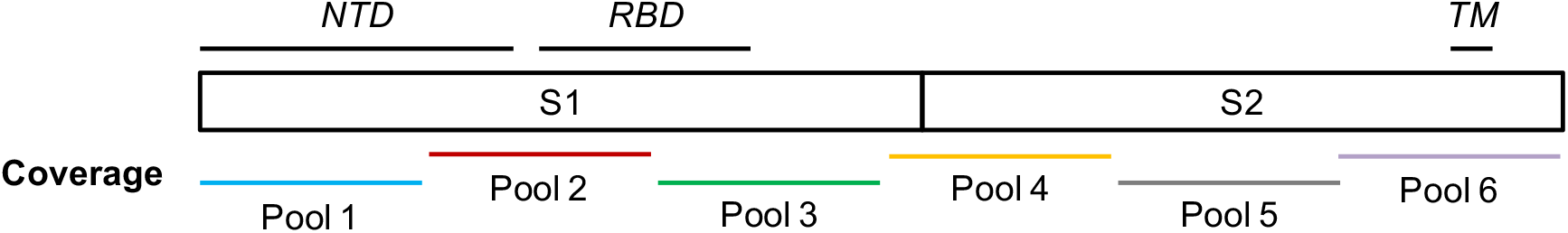
Spike location of the peptide pools used in the cellular assays.

**Figure S2.**
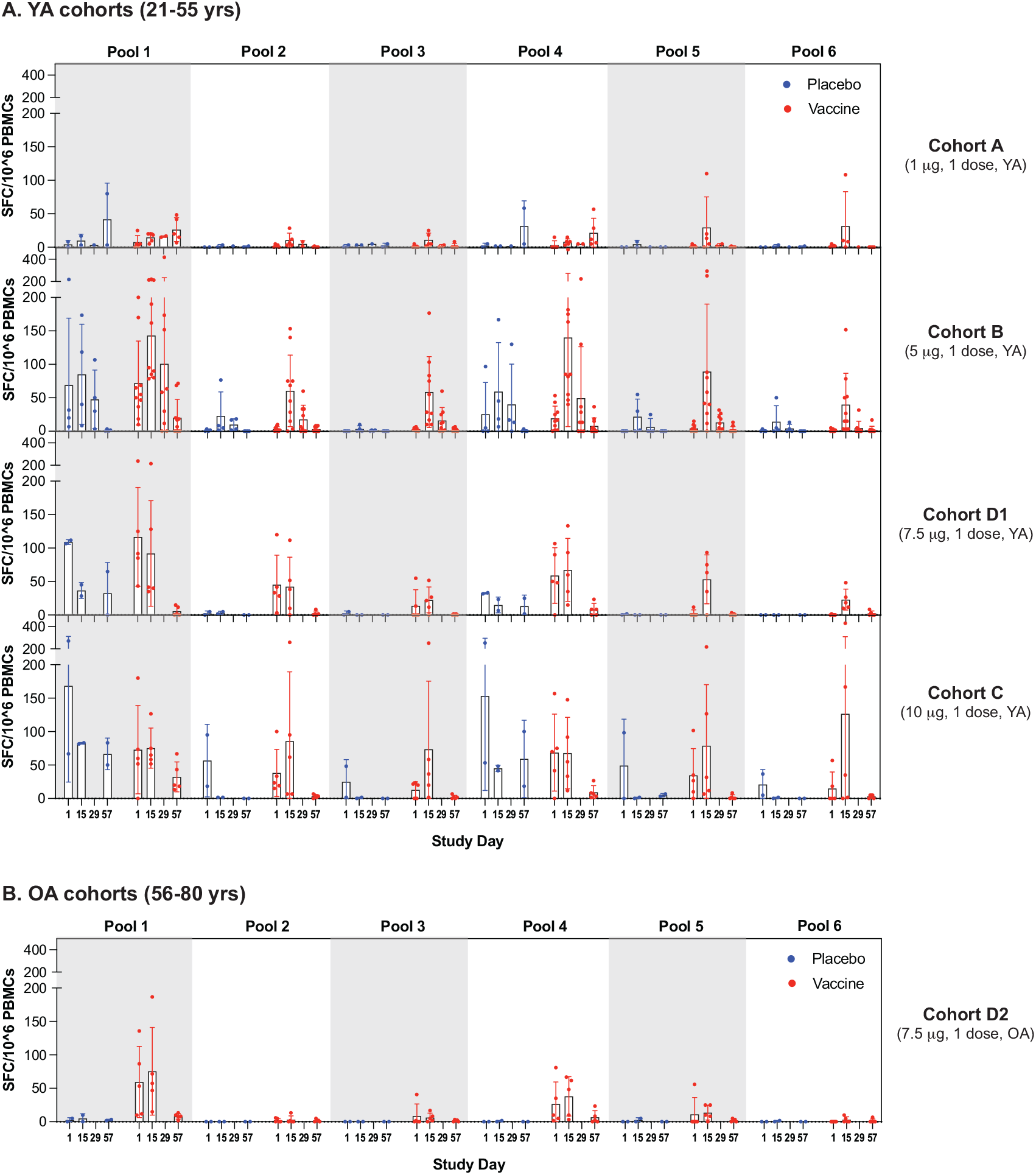
ELISPOT responses by tested peptide pool in 1 dose cohorts. ELISPOT results on study days 1, 15, 29 and/or 57 are presented as spot forming cells (SFC) per 10^6^ PBMCs are displayed for single dose cohorts of younger adults (YA) (Panel A) and older adults (OA) (Panel B). Note that day 29 data is only for Cohorts A and B. Bar graphs display data as mean per group with error bars depicting the standard deviation (SD). Each dot represents an individual participant.

**Figure S3.**
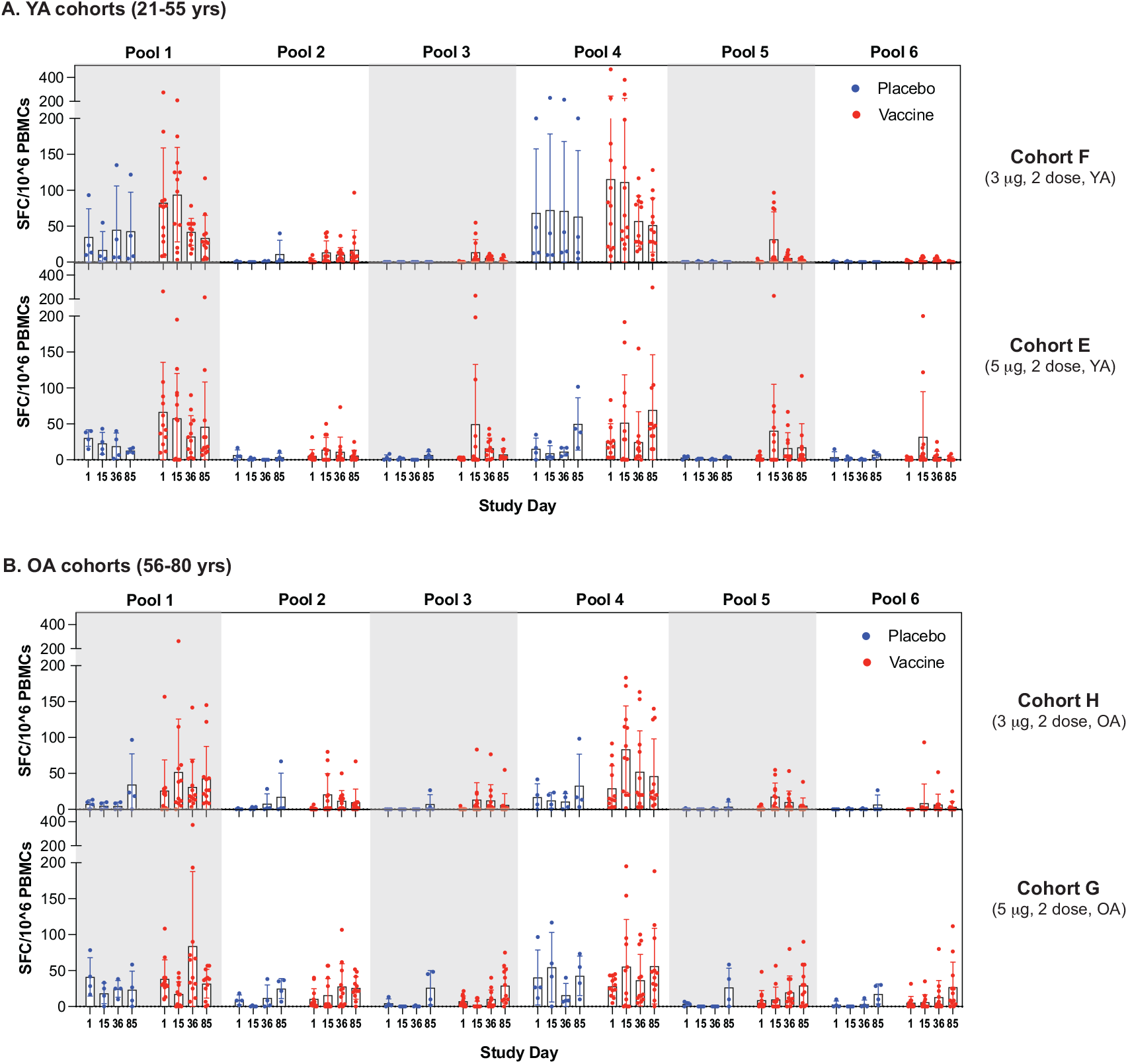
ELISPOT responses by tested peptide pool in 2 dose cohorts. ELISPOT results are presented as spot forming cells (SFC) per 10^6^ PBMCs are displayed for two-dose cohorts of younger adults (YA) (Panel A) and older adults (OA) (Panel B). Bar graphs display data as mean per group with error bars depicting the standard deviation (SD). Each dot represents and individual participant.

**Figure S4.**
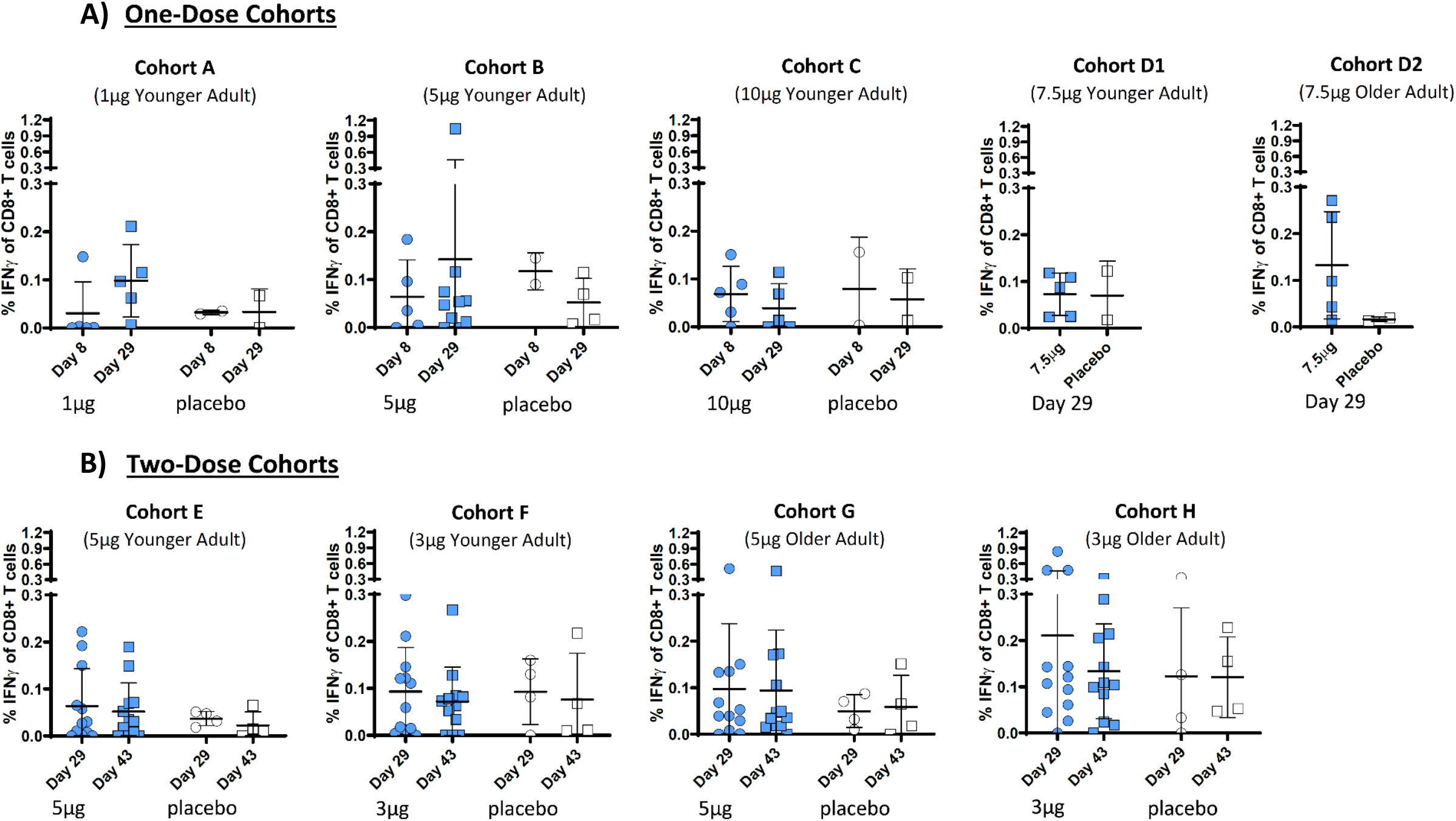
Antigen-specific IFNγ CD8 T cell responses by ICS. Total antigen-specific IFNγ responses in CD8 T cells following stimulation with Spike peptide pools are presented for one-dose (Panel A) and two-dose (Panel B) cohorts. Horizontal bar displays mean per group with error bars depicting the standard deviation (SD). Each dot represents and individual participant. Younger adults 21-55 years; older adults 56-80 years.

**Figure S5.**
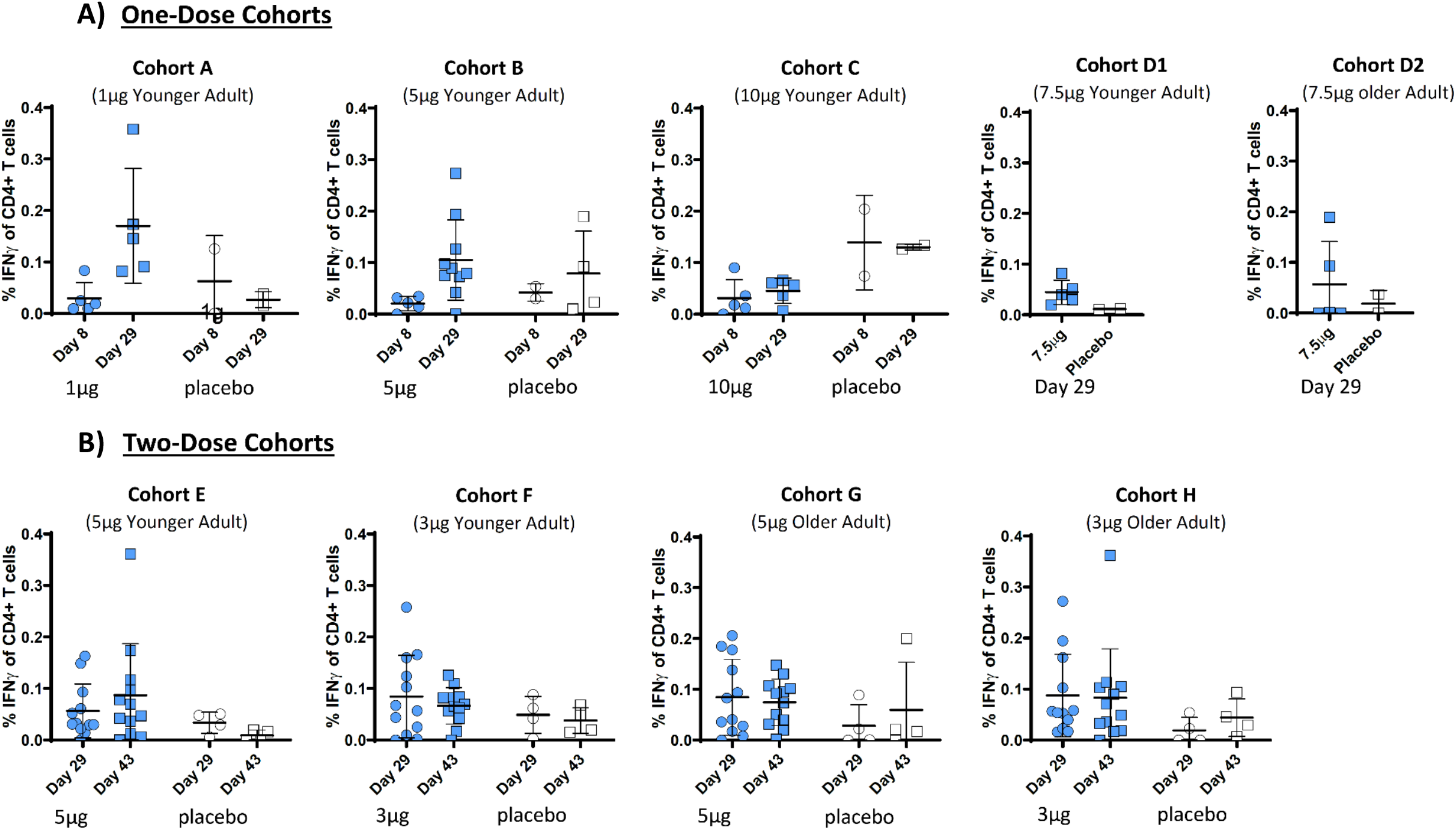
Antigen-specific IFNγ CD4 T cell responses by ICS. Total antigen-specific IFNγ responses in CD4 T cells following stimulation with Spike peptide pools are presented for one-dose (Panel A) and two-dose (Panel B) cohorts. Horizontal bar displays mean per group with error bars depicting the standard deviation (SD). Each dot represents and individual participant. Younger adults 21-55 years; older adults 56-80 years.

**Figure S6.**
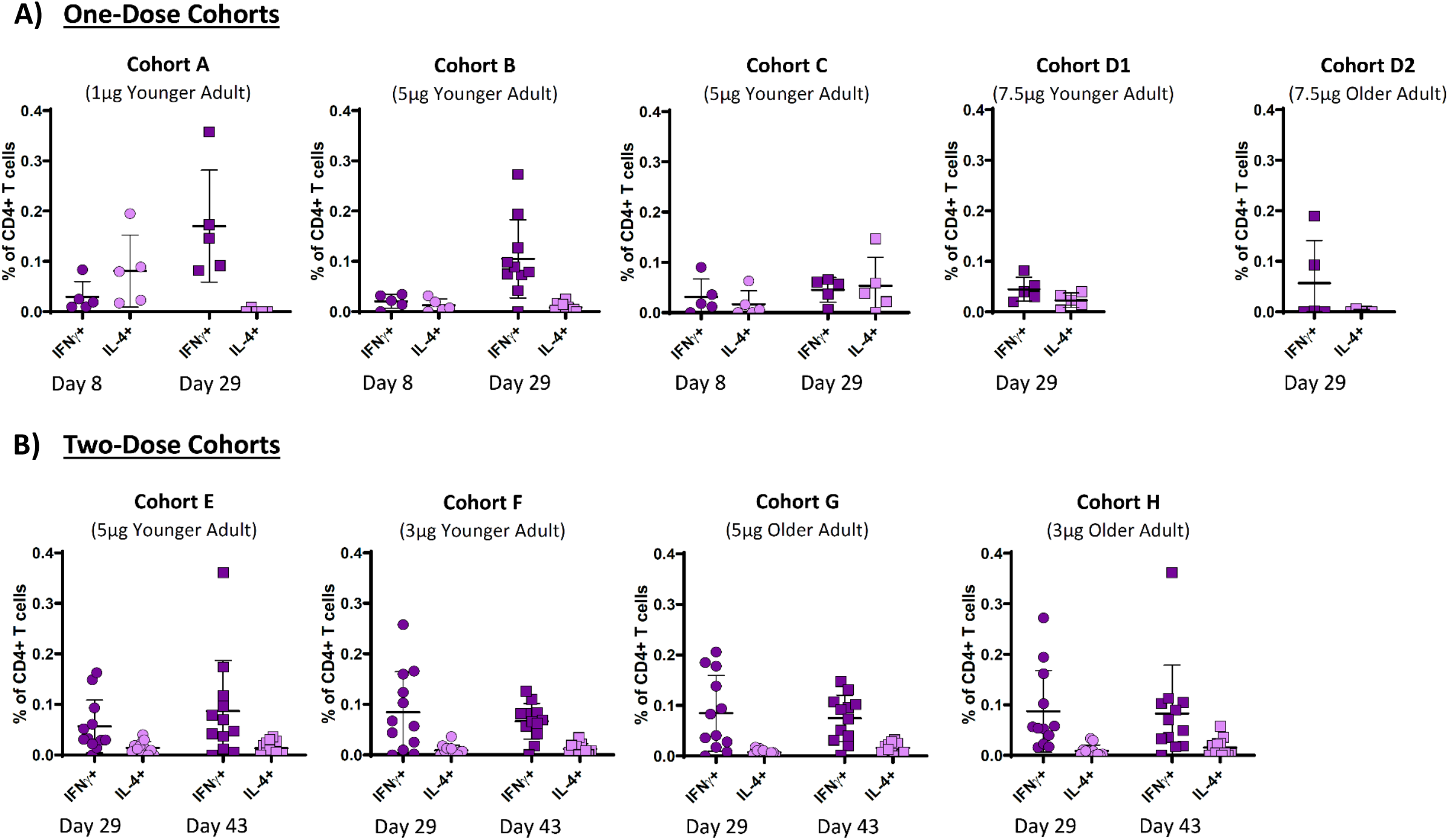
Antigen-specific IFNγ and IL4 CD4 T cell responses by ICS. Total antigen-specific IFNγ and IL4 responses in CD4 T cells following stimulation with Spike peptide pools were measured to assess Th1 responses. Figure presents data for one-dose (Panel A) and two-dose (Panel B) cohorts. Horizontal bar displays mean per group with error bars depicting the standard deviation (SD). Each dot represents and individual participant. Younger adults 21-55 years; older adults 56-80 years.

